# Knowledge Connector: Decision support system for multiomics-based precision oncology

**DOI:** 10.1101/2025.02.23.25322403

**Authors:** Daniel Hübschmann, Simon Kreutzfeldt, Benjamin Roth, Katrin Glocker, Janine Schoop, Lena Oeser, Steffen Hausmann, Christian Koch, Sebastian Uhrig, Jennifer Hüllein, Barbara Hutter, Martina Fröhlich, Christoph E. Heilig, Maria-Veronica Teleanu, Daniel B. Lipka, Irina A. Kerle, Annika Baude, Katja Beck, Christoph Heining, Hanno Glimm, Frank Ückert, Alexander Knurr, Stefan Fröhling, Peter Horak

## Abstract

Precision cancer medicine aims to improve patient outcomes by providing individually tailored recommendations for clinical management based on the evaluation of biological disease profiles in multidisciplinary molecular tumor boards (MTBs). The quality of MTB decisions depends on the comprehensive, reliable, and reproducible interpretation of increasingly complex molecular data. We developed and implemented, as part of a multicenter precision oncology program, the Knowledge Connector (KC), a decision support system that integrates individual patients’ molecular and clinical data with world knowledge to generate and document MTB recommendations. The KC supports data curation, database integration, and discussion based on multiomics data and provides an interface for creating a cross-institutional knowledge base. Furthermore, it extracts relevant biomarker-drug associations and increases the efficacy of data interpretation in a clinically relevant manner by reducing reliance on external sources and optimizing inter-curator concordance. Our results demonstrate that the KC is a versatile tool that supports medical decision-making in MTBs, thus enabling the scalability of precision cancer medicine.

## Introduction

Technologies that enable comprehensive molecular analysis of individual patients’ tumors at high throughput have led to the emergence of precision oncology. This approach is based on identifying, annotating, and evaluating a broad spectrum of molecular biomarkers ranging from single-gene mutations to complex profiles, such as mutational or gene expression signatures, and composite scores combining several of these measures. Such biomarkers aim to capture the predictive, prognostic, and diagnostic value of molecular data to categorize patients into different treatment and outcome groups. Well-established examples from clinical practice include BRAF V600E mutations, which predict response to inhibition of BRAF-MEK signaling in melanoma^1^, or NPM1, FLT3, and CEBPA mutations, which provide prognostic information in acute myeloid leukemia^2^.

Currently, DNA-based sequencing of up to several hundred genes is the most widely used diagnostic tool in precision oncology. However, there is increasing evidence that whole-genome sequencing (WGS), whole-exome sequencing (WES), RNA sequencing (RNA-seq), and probably other omics layers have additional clinical value ^3–7^. Therefore, complex multilayered data must be considered in clinical decision-making, which represents a new challenge in oncology^8^. A critical step to overcome this challenge has been the establishment of dedicated precision oncology workflows that feed into molecular tumor boards (MTBs), where experts from various disciplines, including oncology, medical genetics, bioinformatics, molecular biology, pathology, and others, discuss individual cases. However, expert opinions alone cannot achieve the necessary standardization, reproducibility, and scalability.

The first step in a precision oncology workflow is the molecular characterization of tumor tissue using high-throughput methods, which are becoming increasingly numerous and comprehensive. For example, as part of the MASTER (Molecularly Aided Stratification for Tumor Eradication Research) program of the German Cancer Research Center (DKFZ), the National Center for Tumor Diseases (NCT), and the German Cancer Consortium (DKTK), advanced cancers are routinely subjected to WGS or WES, RNA-seq, and DNA methylation profiling^6,9^. Raw data are processed bioinformatically and forwarded to biomedical curators for functional assessment, followed by the interpretation of specific alterations regarding their oncogenicity and the selection of clinically actionable biomarkers, which are discussed in an interdisciplinary MTB.

The next step is data curation, which aims to obtain a humanly comprehensible set of biomarkers, followed by the preparation, execution, and documentation of MTBs. These workflow components are not standardized, leading to relevant differences between institutions in the practical use of biomarker information and inconsistencies in the reporting of molecular data^10–12^. Consequently, the main bottleneck of MTB workflows is curating and interpreting molecular alterations. This is a complex process in which experts manually search knowledge bases for information on specific variants and corresponding therapeutic options. However, the range of sources and large amount of information make systematic and time-efficient processing difficult, and the content of knowledge bases is heterogeneous^13^. Furthermore, currently available knowledge bases are organized by genes or biomarkers^14,15^. While such a structure covers many applications in precision oncology, there is a growing number of complex biomarkers and alterations at the supragenic level, e.g., polyploidy, large copy number aberrations, and mutational signatures. Another challenge is that evidence in the medical literature usually applies to a specific clinical setting. In precision oncology, an individual case may have some overlap but not an exact match with published evidence.

The ambition to provide clinical care tailored to the unique characteristics of each individual cancer has entailed several new challenges. In addition to considering precision oncology in training programs and the definition of guidelines and standard operating procedures, these mainly include the development of decision support systems, which has become one of the most pressing goals of clinical informatics. These software solutions make molecular tumor profiles accessible and interpretable for data curators and medical professionals by linking them with relevant, accurate, and up-to-date content from knowledge bases and play an increasing role in expediting, standardizing, and ensuring the quality of reporting within and between institutions^16^.

Here, we present the development and implementation of the Knowledge Connector (KC), a web application to support MTB workflows, within the DKFZ/NCT/DKTK MASTER precision oncology network, which uses multiomics for clinical decision-making (https://demo.kc.dkfz.de; code available at https://git.dkfz.de/cto-sudo-pub/demo.kc). We describe how the KC framework facilitates linking individual patients’ molecular profiles with world knowledge, including clinical reasoning based on complex biomarkers and evidence chains. We further introduce the concept of blocks of clinical knowledge (BoCKs) to map evidence to individual biomarkers and explain that the KC also serves as an interface for creating a cross-institutional knowledge base, the BoCKbase.

## Results

### Development of the KC as a decision support system for MTBs

The identification and interpretation of molecular biomarkers is a multistep process^17^. To support several of these steps, particularly biomarker annotation, oncogenicity classification, clinical interpretation, and clinical decision-making (**Fig. 1a** and **Extended Fig. 1**), we developed the KC. The KC (https://demo.kc.dkfz.de; code available at https://git.dkfz.de/cto-sudo-pub/demo.kc) is a web-based, European Union General Data Protection Regulation-compliant system that allows protected access to pseudonymized clinical and molecular data and supports the fast, accurate, and complete execution of MTB workflows using comprehensive molecular data, e.g., from WGS and RNA-seq. The home screen displays meta-information about the tumor sample and patient and condensed clinical data. In a curation and interpretation process guided by a navigation bar at the top of the page, users can select different views ranging from analyzing integrated biomarkers and biomarker scores, inspecting individual biomarkers, evaluating pharmacogenomic information, curating and suggesting treatment recommendations, to presenting the data in the MTB (**Fig. 1b**). The KC thus combines essential clinical and complex molecular data in an intuitive and comprehensive format.

**Fig. 1:**
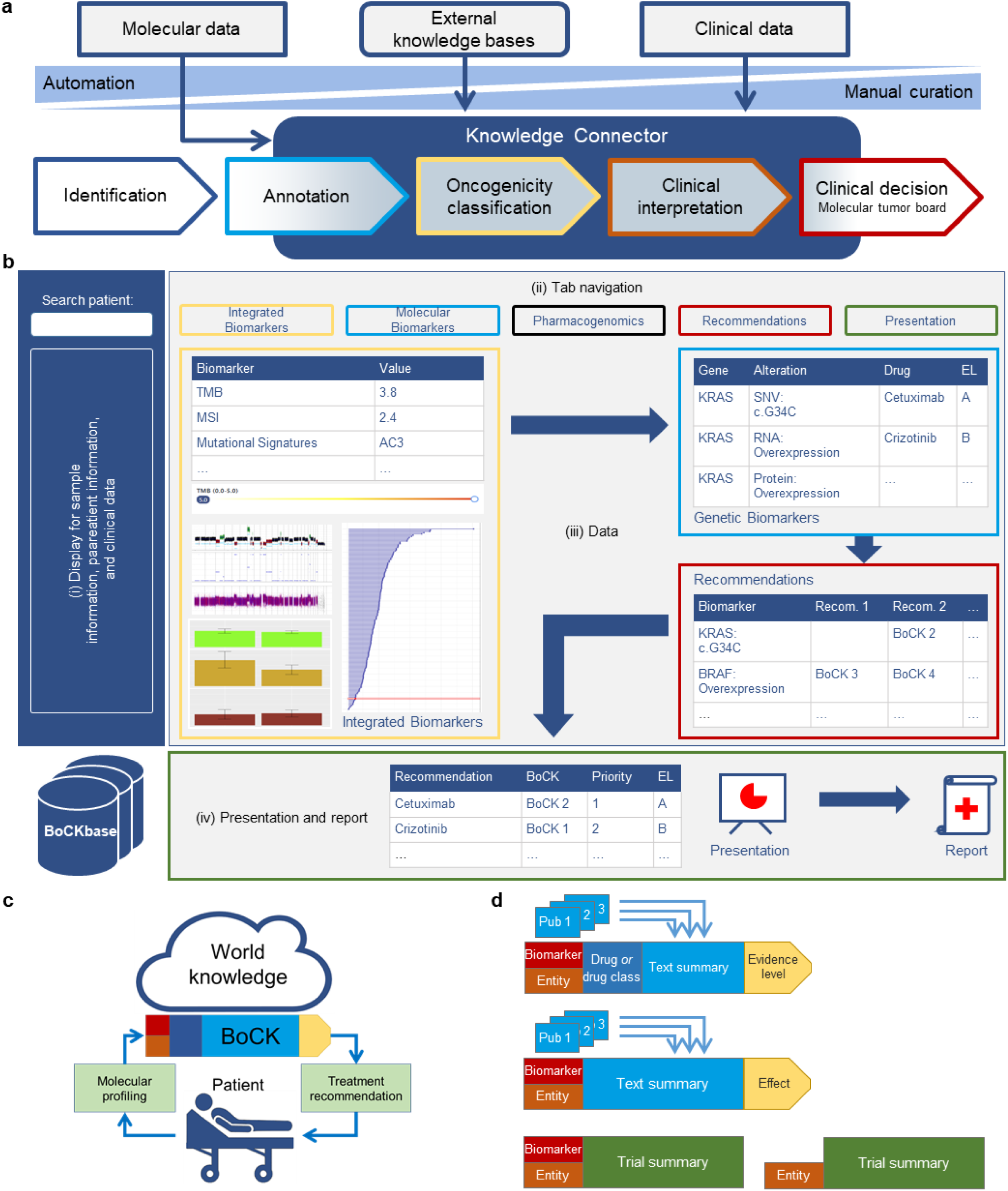
Scope and functionality of the KC. **a,** Precision oncology workflow steps supported by the KC. **b,** Schematic overview of the KC including (i) meta-information on the tumor sample and the patient and clinical data, (ii) the navigation bar, (iii) content selected via the navigation bar, and (iv) the presentation step consisting of a series of slides and concluding with an editable MTB recommendation and prioritization table and a downloadable report. **c,** Link between molecular and clinical data from individual patients with world knowledge via BoCKs. **d,** Examples of BoCKs. Top, BoCK combining a biomarker (e.g., a specific gene mutation), an entity, a drug or drug class, and published evidence, resulting in a predictive statement with a specific evidence level (EL). Middle, BoCK combining a biomarker, an entity, and published evidence, resulting in oncogenicity classification. Bottom, BoCK combining a biomarker, an entity, and published evidence, identifying eligibility for a specific clinical trial. AC, COSMIC (Catalogue Of Somatic Mutations In Cancer) mutational signatures version 2; MSI, microsatellite instability; Pub, publication; Recom., recommendation; SNV, single-nucleotide variant.

Specifically, the main view of the KC consists of three sections (**Fig. 1b**):

1. To the left of the screen, a vertical bar displays meta-information about the tumor sample and clinical data, such as patient identifier, date of birth, diagnosis, or relevant previous treatments.
2. At the top of the screen, a navigation bar enables essential steps in MTB preparation:

a. *Integrated biomarkers*: Information and underlying evidence on complex and composite biomarkers that exceed the single-gene level, e.g., tumor mutational burden (TMB), mutational signatures, or quantitative measures of genomic instability
b. *Molecular biomarkers*: Information and underlying evidence on clinically actionable biomarkers at the single-gene level
c. *Pharmacogenomic information*: Recommendations of the Clinical Pharmacogenetics Implementation Consortium for selected pharmacogenes
d. *Recommendations*: Grouping and enrichment of information selected in steps a. and b., assignment of treatments to biomarkers, and assessment of molecular evidence levels (mEL)^18^
e. *Presentation*: Compilation of the selected biomarkers and recommended treatments for the MTB, final assignment of mEL, and ranking of recommendations
3. The largest part of the screen is a frame for displaying the molecular data, enriched with world knowledge selected via the navigation bar. This adjustable frame may contain tables, graphics, text, editable text fields, pulldown menus, etc.

For the annotation of relevant molecular findings, comprehensive resources, including, but not limited to, Ensembl^19^, OncoKB^20^, CIViC^15,21^, Reactome^22^, and JAX-CKB^23^, or individual evidence items, e.g., from PubMed, are used. To optimize integrating comprehensive molecular and clinical data from individual patients with world knowledge (**Fig. 1c**), we have introduced the concept of BoCKs. These aggregated statements cite diagnostic, prognostic, predictive, or clinical trial evidence for a particular biomarker consistent with a patient’s tumor entity (**Fig. 1d** and **Extended Data Fig. 2a**). Clinical decisions often consider indirect matches, e.g., when a particular molecular alteration is oncogenic (**Fig. 1d** and **Extended Data Fig. 2b**) or predictive in another tumor entity, or by extending the search for a specific drug to other compounds with similar pharmacodynamic effects. BoCKs can accommodate this type of reasoning and also include information on biomarker-stratified clinical trials (**Fig. 1d** and **Extended Data Fig. 2c**). All curated BoCKs are stored in a database, the BoCKbase, and are reusable for future curation, generating a growing resource for molecularly informed clinical decision-making at the individual-patient level.

### Use of the KC for MTB preparation

To illustrate the KC’s functionality and unique features in assessing comprehensive, multidimensional tumor profiles, we selected cases from the MASTER program^6,9^ with biomarkers not usually detected by standard technologies, i.e., DNA-based panel sequencing.

**Homologous recombination deficiency.** Homologous recombination deficiency (HRD) can leave various imprints on the genomes of cancer cells, including specific mutational signatures, such as single-base substitution signature 3 (ref. 24-26), and quantifiable manifestations of genomic instability^27–29^. Such imprints, which predict sensitivity to poly(ADP-ribose) polymerase (PARP) inhibitors or platinum-based chemotherapy, are often detected in the absence of “classical” loss-of-function mutations in genes associated with homologous recombination, such as BRCA1/2. The KC provides an integrated and intuitive display of the two genomic imprints of HRD, accessible via the “Integrated Biomarkers” tab. Genomic instability is indicated by three parameters (**Fig. 2a**): the loss of heterozygosity (LOH)-HRD score^27^, the number of large-scale state transitions (LSTs)^29^, and the amount of telomeric allelic imbalance (TAI)^28^. Exposure to mutational signatures computed with YAPSA^25^ is displayed as stacked bar plots. **Fig. 2b** shows an HRD phenotype in a patient with undifferentiated thyroid carcinoma. This unexpected finding in this entity illustrates the functionality of the KC as a visualization and decision support tool and prompted screening for participation in a molecularly stratified clinical trial of combination treatment containing the PARP inhibitor olaparib (ClinicalTrials.gov: NCT03127215).

**Fig. 2:**
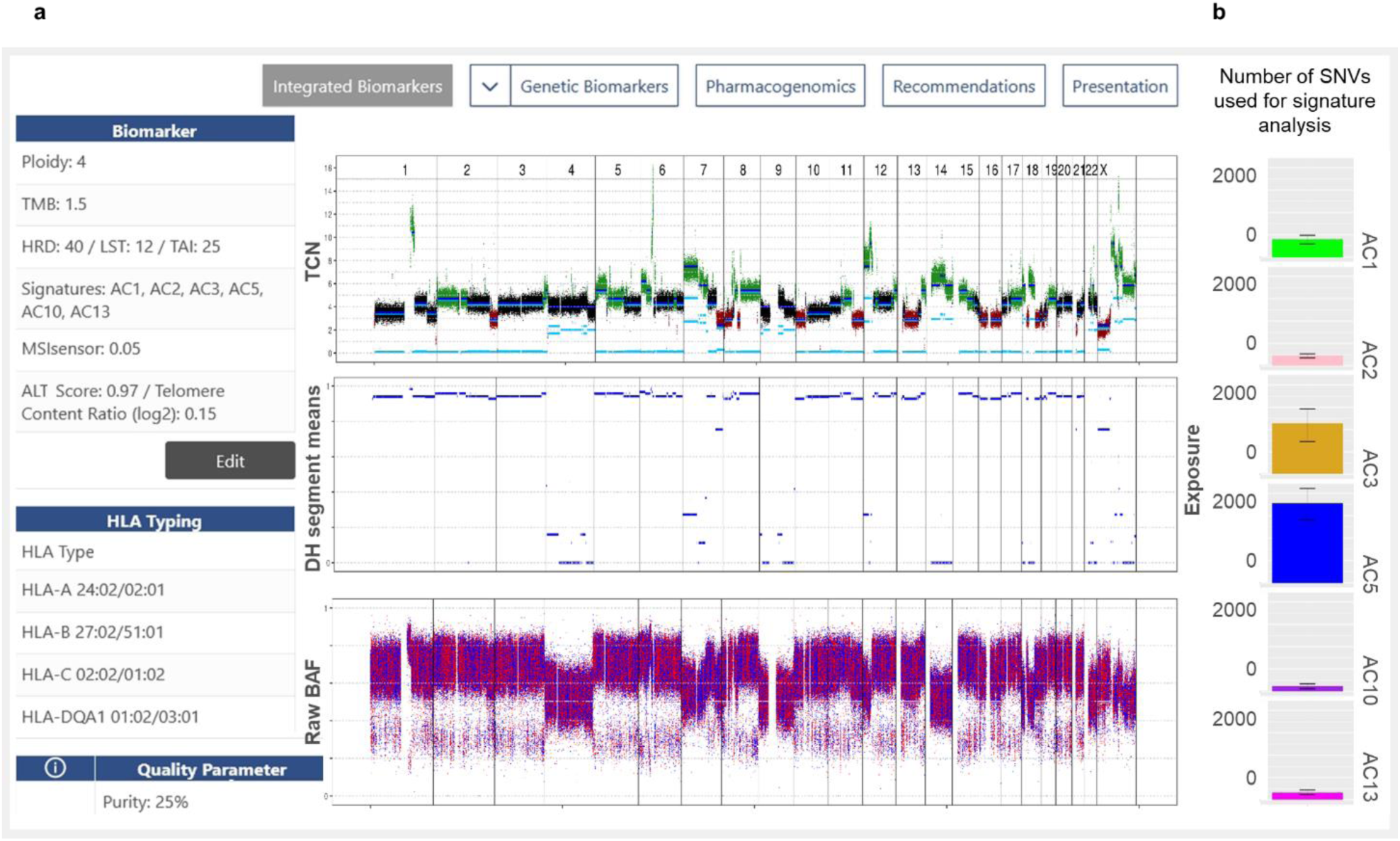
KC display of HRD and mutational signatures. **a,** Left, complex biomarkers, displayed via selection of the “Integrated Biomarkers” tab in the navigation bar, with numerical values for HRD, LSTs, and TAI to quantify genomic instability. Display frame on the right, visualization of total copy number (TCN), decrease of heterozygosity (DH), and B-allele frequency (BAF). **b,** Visualization of exposures to mutational signatures as alternative content of the display frame. ALT, alternative lengthening of telomeres; AC, mutational signature.

**Gene fusions.** The KC also allows the visualization and reporting of RNA-seq-based gene fusion calls of the software Arriba^30^. The “Genetic Biomarkers” view of the KC integrates all alteration types affecting a gene of interest, e.g., BRAF in a patient with parotid gland carcinoma harboring a previously unknown AGK::BRAF fusion that resulted from a genomic inversion reported as structural variant (SV; **Fig. 3a**). Transcriptomic analysis confirmed the fusion of AGK as the N-terminal and BRAF as the C-terminal partner. The KC provides a graphical representation of several features of the fusion derived from Arriba, i.e., the location of the breakpoints relative to the coding sequence, the exons involved, the orientation (sense or antisense) of the fusion product (**Fig. 3b**), the chromosomal position (**Fig. 3c**), and the preserved functional domains of the resulting protein (**Fig. 3d**). The AGK::BRAF fusion involved BRAF from exon 8, including the kinase domain from exon 11, and retained the open reading frame. Therefore, the fusion was considered to activate the MAPK pathway. This evaluation was supported by gene-level information from internal and external knowledge bases, summarized in BoCKs with different mEL. Preclinical work (mEL 3) showed an effect of the MEK1/2 inhibitor trametinib in AGK::BRAF-positive melanoma models^31^. In addition, AGK::BRAF fusions have been detected in cancer patients, and response to MEK inhibition has been observed in individual cases (mEL 2C)^32–34^. For example, a patient with AGK::BRAF-driven melanoma showed resistance to vemurafenib but sensitivity to sorafenib in vitro and subsequently a durable response to sorafenib in vivo^32^, and a patient with AGK::BRAF-positive myoepithelial carcinoma responded to cobimetinib^34^. Given these BoCKs, one MTB recommendation was sorafenib and/or a MEK inhibitor. Another example of fusion visualization in the KC is a patient with melanoma and a TRIM33::BRAF fusion between two amplified genomic regions (TCN of 8 in an inferred diploid genome; **Extended Data Fig. 3a**). The fusion product included exons 1–11 of TRIM33 and BRAF from exon 9 (**Extended Data Fig. 3b**). This fusion was one of several detected (**Extended Data Fig. 3c**), and its clinical significance was readily identified using the KC, as the predicted chimeric protein included the kinase domain and retained the BRAF open reading frame (**Extended Data Fig. 3d**). Based on these findings and supported by several BoCKs, pan-RAF and/or MEK inhibition were recommended by the MTB.

**Fig. 3:**
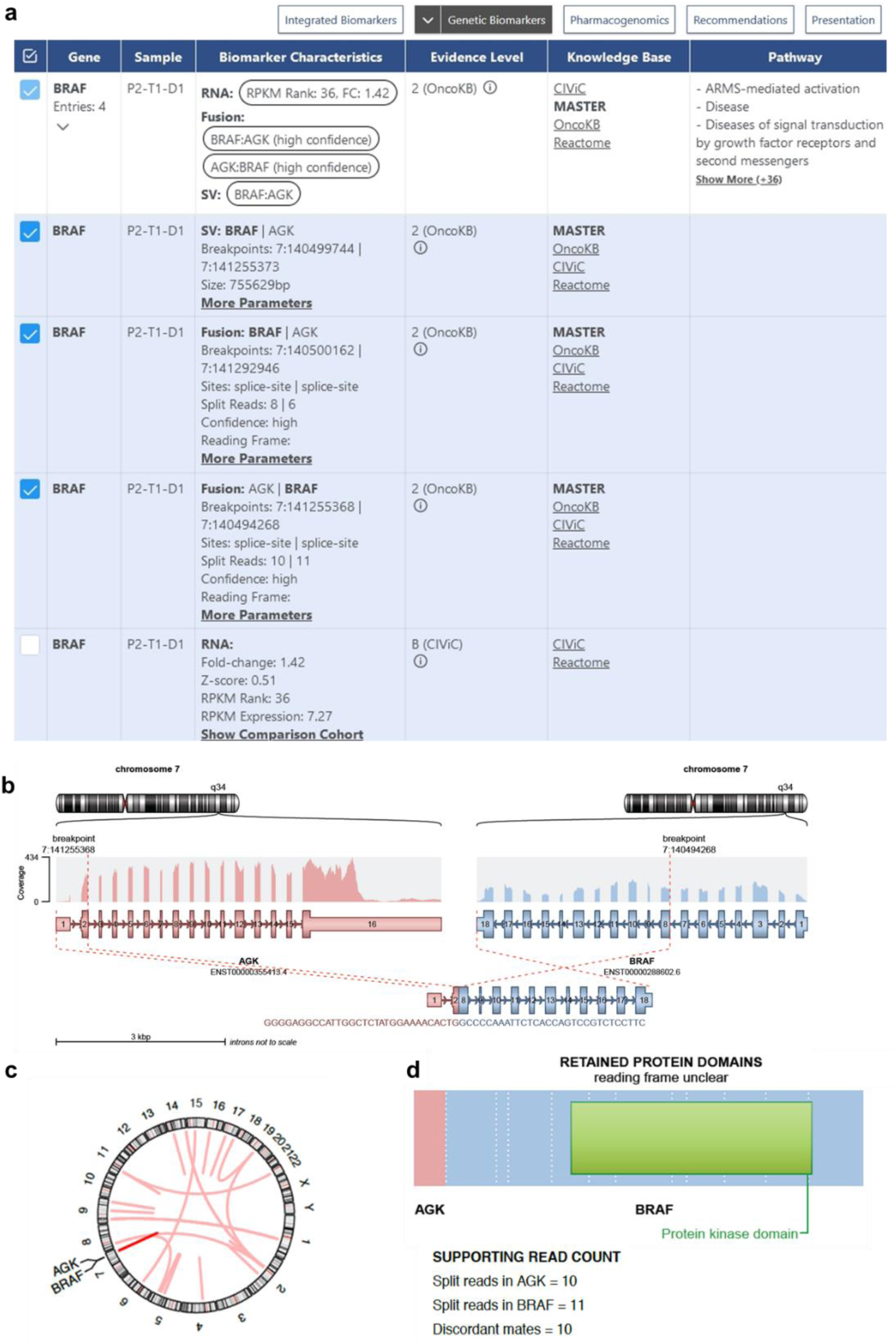
KC display of an AGK::BRAF fusion. **a,** Screenshot of the “Genetic Biomarkers” panel illustrating the congruent detection of a genomic SV and an overexpressed fusion transcript. **b,** Visualization of the AGK::BRAF fusion event by the fusion detection tool Arriba. **c,** Chromosomal position of the AGK::BRAF fusion. **d,** Domain structure of the resulting AGK::BRAF fusion protein. FC, fold change; kbp, kilobase pair; RPKM, reads per kilobase of transcript per million mapped reads.

**Aberrant gene expression.** The KC also allows the integration and exploratory analysis of data generated with emerging technologies such as transcriptomic, epigenomic, and proteomic profiling. For example, RNA-seq is becoming increasingly important for clinical decision-making as, beyond gene fusion detection, it allows the quantification of mRNA expression of structurally intact genes and their isoforms, whose protein products are targeted by a growing number of therapies. **Fig. 4a** shows the case of a patient with a mediastinal non-seminomatous germ cell tumor characterized by overexpression of CLDN6 (2,193-fold, rank 1 of the reference subcohort). Based on this finding, treatment with CLDN6-directed chimeric antigen receptor T cells was recommended (mEL 1C)^35,36^ as part of a clinical trial (NCT04503278). Another example is a patient with advanced Ewing sarcoma and a 210-fold (rank 2) increase in mRNA expression of the PRAME cancer testis antigen (**Fig. 4b**). PRAME is a potential target for the development of cellular immunotherapies in sarcoma (mEL 4)^37^, while in medulloblastoma, preclinical studies have shown the efficacy of adoptive immunotherapy with PRAME-specific T cells (mEL 3)^38^. Analogous to the example of CLDN6, at the time of the MTB, a matching clinical trial (NCT03686124) was recruiting, whose eligibility criteria included HLA A*02:01 positivity, which was confirmed by *in silico* HLA typing from DNA sequencing data and validation using orthogonal methods. The KC also displays such information to the MTB, enabling rapid evaluation of a patient’s potential eligibility for clinical trials.

**Fig. 4:**
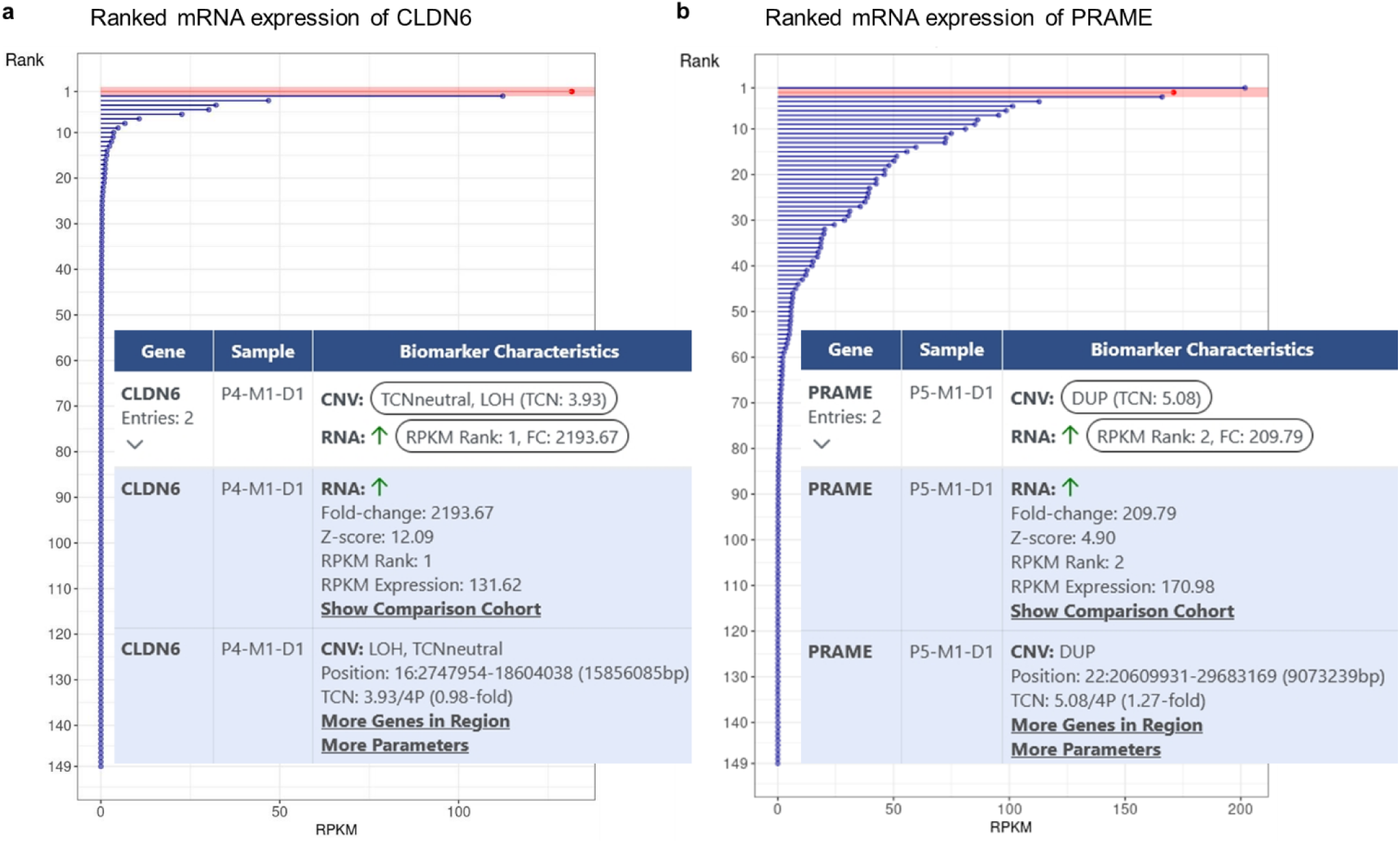
KC display of expression biomarkers. **a,** CLDN6 mRNA expression. The expression rank of the patient described is indicated in red. Inset, screenshot of the “Genetic Biomarkers” panel illustrating CLDN6 overexpression. **b,** PRAME mRNA expression. The expression rank of the patient described is indicated in red. Inset, screenshot of the “Genetic Biomarkers” panel illustrating PRAME overexpression and a DNA copy number gain in the corresponding genomic region. CNV, copy number variant; DUP, duplication; TCNneutral, copy number neutral LOH; 4P, inferred ploidy of 4.

### Curation of clinical biomarker associations

Once the relevant molecular alterations are identified, the curation process proceeds to their assignment of recommendations and evidence defined in BoCKs. In the corresponding KC view, all predictive, prognostic, and diagnostic biomarkers are displayed in a compact form, and the clinical consequences, including therapeutic intervention, genetic counseling, or pathologic reevaluation, can be determined. If one or several BoCKs show biomarker-drug associations, multiple drugs and/or drug classes^39^ (**Extended Data Fig. 4a**) and clinical trial BoCKs (**Extended Data Fig. 4b**) can be assigned to the recommendation. The KC aggregates information from available databases and automatically matches it to the selected biomarker. However, since information obtained from different knowledge bases is heterogeneous and automatic matching is unreliable^13^, subsequent human curation, correction, and confirmation are required. **Fig. 5** depicts the process of curating an evidence-based MTB recommendation. Based on the selection of relevant biomarkers, BoCKs are used to formulate one or more recommendations, which are assigned a cumulative, preliminary molecular evidence. In the second step, an analogous assignment is made for clinical trials, which can only be partially automated and must consider patient eligibility and trial availability. In the final step, the curated recommendations for clinical management and trial enrollment are presented to and discussed by the MTB. As a visualization and decision support system, the KC interacts with a continuously growing, manually curated database for BoCKs, the BoCKbase. The BoCKbase can be made accessible to multiple users and institutions, making it a user-maintained, quality-controlled resource that evolves and leads to a community consensus through the use of the KC in MTB preparation.

**Fig. 5:**
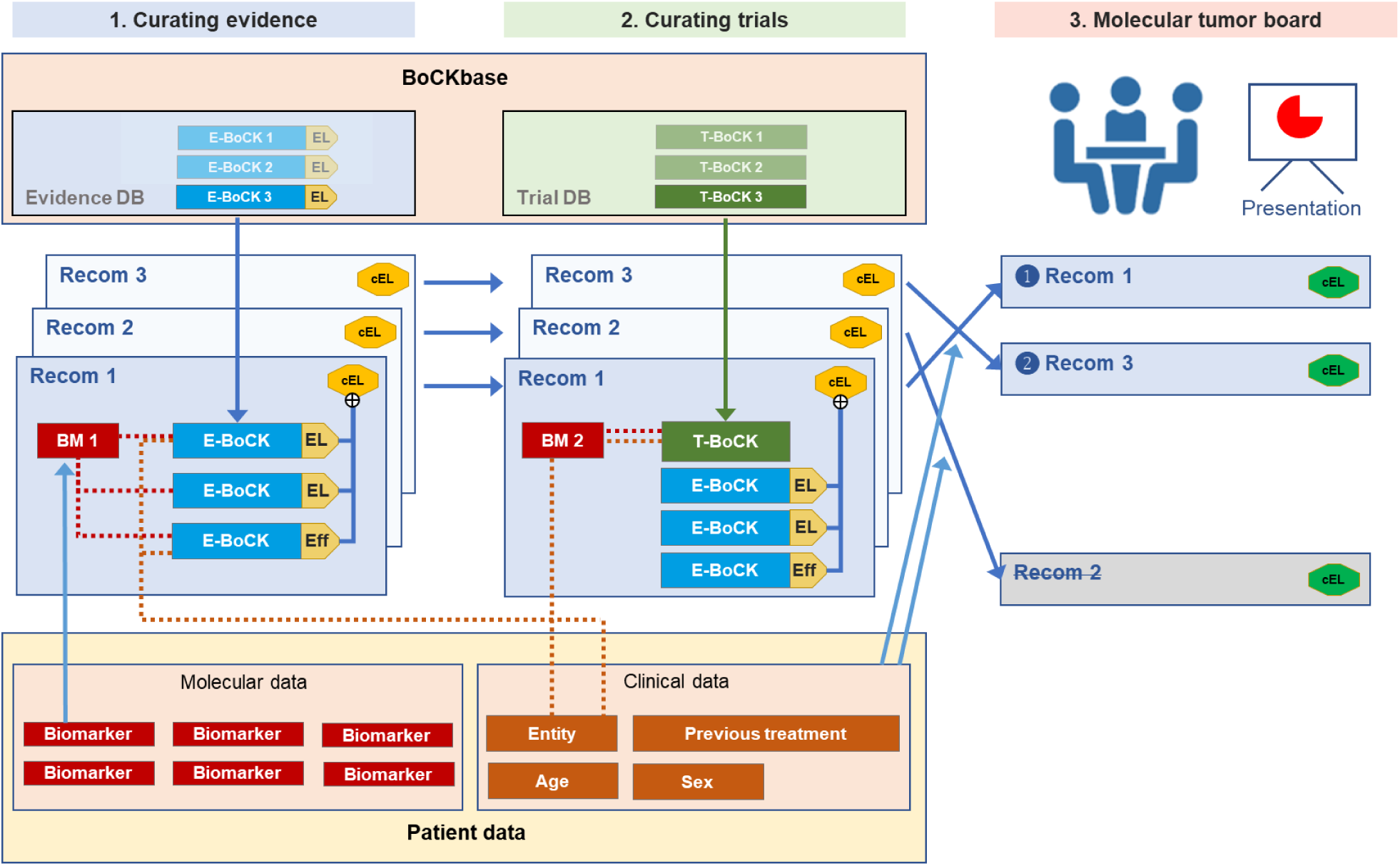
Curation of evidence-based MTB recommendations. Curating recommendations for individual patients in precision oncology involves three steps. For each recommendation, clinical and molecular data (bottom) are linked to BoCKs (top). The first step is to link to medical interventions, followed by clinical trials. The third step comprises the presentation and clinical decision-making, including prioritization of recommendations, in the MTB. BM, biomarker; cEL, clinical evidence level; DB, database; E-BoCK, evidence block of clinical knowledge; Eff, effect; T-BoCK, clinical trial block of clinical knowledge.

### Real-world application of the KC

The KC has been in productive use at NCT Heidelberg since February 2022 and has since been implemented at several other comprehensive cancer centers in Germany. **Fig. 6a** shows the KC-based molecular data curation and discussions in the MTB of the first 268 cases, including the distributions of the number of recommendations, actionable biomarkers, BoCKs, and clinical trials assigned per patient. At least one recommendation for clinical intervention was made in 243 of 268 patients (90.7%; median, 3; range, 1–7). A suitable molecularly stratified clinical trial was identified in 151 cases (56.3%). The high yield of biomarkers per patient (median, 5; range, 1–16) reflects the breadth of information contained in comprehensive molecular profiles, and the assignment of up to 20 BoCKs per patient (median, 7) demonstrates the utility of this condensed format for biomarker curation and information transfer to the MTB. The number of newly generated BoCKs stored in the BoCKbase is steadily increasing (**Fig. 6b**). As expected, it was initially proportional to the number of cases processed cases but decoupled after approximately four months, demonstrating the reuse of previously stored BoCKs. This decoupling reflects the potential of BoCKs developed and maintained as part of biomedical curation in a clinical setting to create a knowledge base that increases the efficiency and scalability of precision oncology workflows. The molecular alterations used for clinical recommendations include a broad spectrum of single-gene and complex biomarkers (**Fig. 6c**). Most DNA-based single-gene biomarkers are SNVs and somatic CNVs, of which those affecting CDKN2A, PTEN, and PIK3CA are most commonly used for MTB decisions (**Fig. 6d**). At the RNA level, aberrant expression of individual genes dominates (**Fig. 6c**), often prompting the recommendation of targeted therapies, e.g., against CDH17, FOLR1, or TACSTD2 (**Fig. 6d**). Complex biomarkers comprise mutational signatures, measures of genomic instability, and TMB estimates (**Fig. 6c**).

**Fig. 6:**
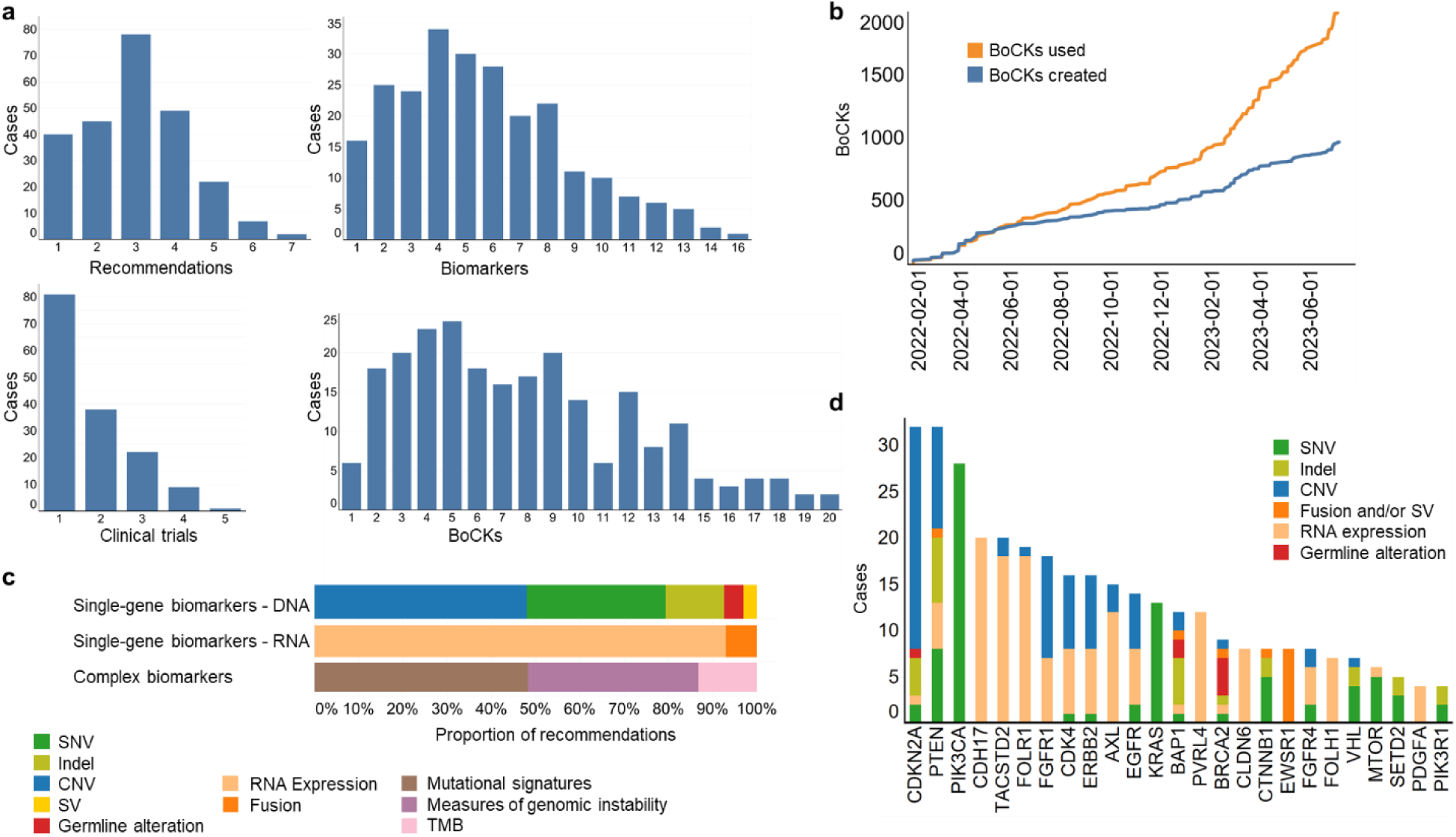
Overview of 268 MTB cases curated using the KC. **a,** Number of recommendations (total, n=243), biomarkers (total, n=241), clinical trials (total, n=151), and BoCKs (total, n=235) per MTB case. **b,** Cumulative number of BoCKs created in the BoCKbase and used for clinical decision-making over time. **c,** Proportion of single-gene and complex biomarkers. **d,** 20 most frequently used biomarker genes and their molecular alterations. Indel, small insertion/deletion.

## Discussion

We here describe the KC, a data integration, curation, visualization, and decision support platform for precision oncology workflows. By providing a flexible and user-friendly interface, the KC reduces manual workload, thereby facilitating clinical decision-making in MTBs and increasing their scalability.

Specific tasks in the KC workflow require expertise from different domains and thus the involvement of multiple people in the multistep process of curating molecular data for decision-making in MTBs, which includes variant calling and the identification, annotation, functional classification, and clinical evaluation of biomarkers. While variant calling and the identification and annotation of biomarkers can be standardized and automated, functional and clinical evaluation remain manual tasks. Evidence for the relevance of a specific biomarker relies, with few exceptions, on unregulated external knowledge bases and needs to be organized and presented to the MTB in a concise and patient-centric manner. Clinical guidelines and standard operating procedures for these tasks are being developed but are highly heterogeneous across institutions and countries^40^. The KC, a software tool for extracting actionable information from multilayered molecular cancer profiles, supports several steps of this process by providing annotation and structure for the functional and clinical assessment of biomarkers, considering the specifics of the diagnostic technologies applied. It aggregates information from multiple sources and provides biomedical curators with all necessary algorithmic functions for sorting, filtering, and the functional assessment of biomarkers using various views to optimally inform molecularly guided treatment decisions. In addition, bioinformatic quality controls for the approval of biomarkers are part of the KC.

Of note is that the KC supports recommendations for clinical management based on single-gene as well as complex and composite biomarkers. The underlying data structures are flexible, allowing for rapid integration and development of new, user-defined composite biomarkers, whose relevance increases as molecular testing becomes more comprehensive. The KC decision support system can already accommodate biomarkers based on a wide range of molecular information layers, i.e., the genome (including germline variants associated with hereditary cancer predisposition or pharmacogenomic risk), transcriptome, DNA methylome, and phosphoproteome, enabling multiomics integration at the patient level. Ongoing developments focus on user role development and incorporating data generated by additional tumor profiling methods, such as digital pathology approaches and functional drug screens, or new bioinformatics tools.

Another essential feature of the KC is its database functionality, which is based on the concept of BoCKs. While the BoCKbase complements rather than replaces external knowledge bases, it enables the merging of evidence items from different sources based on diverse data models, thus facilitating the reuse and storage of BoCKs. Another strength of BoCKs is the generation of content for medical reports in the working language of the respective MTB, facilitating implementation in non-English speaking countries. In addition, the BoCKbase can be implemented and maintained across institutions, thereby leveraging the collective expertise of precision oncology networks.

There are several academic and commercial efforts aimed at creating cross-institutional MTB platforms^16,41–43^. The KC has two distinct features that make it particularly suited to address the challenge of processing multilayered molecular profiles: (a) by focusing on the curation of biomarkers and their mapping to drugs without committing to specific bioinformatic functions, it is technology-agnostic and can be adapted to any portfolio of molecular profiling methods; (b) by balancing the level of automation to increase productivity and not forcing algorithmic decisions, the KC is open to ongoing developments and the implementation of standard operating procedures for biomarker annotation and functional classification. For precision oncology to reach its full potential, patients’ molecular profiles must also be linked to information from electronic health records (EHRs) and previous MTB evaluations to create a sustainable resource of biomarker profiles, treatment recommendations, and, ideally, patient outcomes. Sharing such data collections across institutions will also help standardize MTB recommendations and enable less experienced physicians and, thus, patients to benefit from the expert knowledge generated.

In summary, we have developed a novel point-of-care software solution that extracts clinically relevant biomarker-drug associations from comprehensive, multilayered molecular cancer profiles, increases the efficiency of data curation and interpretation, and links molecular and clinical data at the individual-patient level. Integrating the KC into precision oncology workflows will substantially increase their quality, standardization, and throughput, which is essential for the widespread adoption of molecularly informed cancer medicine in an increasing number of patients.

## Materials and methods

### Data and Code Availability

The Knowledge Connector (KC) is a web application to support MTB workflows, which use multiomics for clinical decision-making. A demo instance is available at https://demo.kc.dkfz.de. In oder to obey to requirements of protection of patient information, this demo istance contains only mock patients. The data of these patients is available at https://demo.kc.dkfz.de. Of note, as this work does not present a clinical trial, aspects of randomization, control group, blinding or power analysis are not applicable. The code is available at https://git.dkfz.de/cto-sudo-pub/demo.kc.

### Patient-related data

Patients’ medical histories are obtained from EHRs based on data points defined by medical oncologists, bioinformaticians, and medical informaticians, i.e., patient and biospecimen identifiers, date of birth, sex, diagnoses, topologies and morphologies of the primary tumor and metastases, therapies administered, and response and survival data. Molecular data from, e.g., WGS or WES and RNA-seq, are obtained as processed data files from the bioinformatics workflow. Raw sequencing data are analyzed by a set of pipelines operated by the One Touch Pipeline, an automated workflow and data management system^44^. Briefly, WGS and WES data are aligned with a workflow based on bwa^45^, and variant calling is performed using mpileup for germline and somatic SNVs, platypus for germline and somatic indels, ACEseq^46^ for somatic CNVs from WGS, cnvkit^47^ (RRID:SCR_021917) for somatic CNVs from WES, and SOPHIA for germline and somatic SVs. RNA-seq data are aligned with STAR^48^ (RRID:SCR_004463), transcripts are quantified using featureCounts (RRID:SCR_012919), and gene fusions are called by Arriba^30^ (RRID:SCR_025854). All bioinformatics pipelines have been benchmarked and described previously^6,49^.

### Communication between the KC and other software

The KC uses as input a patient’s clinical and molecular data on the one hand and world knowledge on the other. Each interface is implemented as REST API. Clinical and molecular data are transferred (**Extended Data Fig. 1**) via the KC Data Pool to the KC Database, which consists of a PosgreSQL database and a data model specified for the KC (**Supplementary Fig. 1** and **Supplementary Table 1–24**). The individual elements of the data model do not enforce a controlled vocabulary, providing a flexible interface to accommodate different ontologies and classifications. Clinical data from various EHR systems can be integrated through transformation into the existing data model of the KC. Additional bioinformatics pipelines can be integrated by adding the appropriate tables to the existing data model. Queries with patient-related clinical and molecular data can be sent to the KC Database via one of the REST APIs to retrieve the content required for clinical decision-making in the MTB. To link clinical and molecular data with world knowledge, the KC performs fully or semi-automated weekly downloads of five external knowledge bases (Ensembl (RRID:SCR_002344), OncoKB (RRID:SCR_014782), CIViC, Reactome (RRID:SCR_003485), JAX-CKB; **Supplementary Table 25**), and MTB-relevant data are transformed into a relational database model and stored in the shared BoCKbase. Before that, comparable information from the different knowledge bases is harmonized. For this purpose, data fields are combined, and uniform terminology is applied while ensuring that the original data record can always be traced back to the respective knowledge base. Like the KC Data Pool, the BoCKbase consists of a PosgreSQL database, and the harmonization and transfer of data are done via SQL.

### Enrichment of patient-related data

Patient-related data in the KC Data Pool are enriched overnight with data from the BoCKbase and transferred to a third PosgreSQL database, the KC Patient Database, for display in the KC frontend. Application interfaces (REST API) are implemented for communication and data exchange between the databases, which are also used by the KC frontend to retrieve the data to be displayed from the KC Patient Database and the BoCKbase (**Extended Data Fig. 4**). Various SQL methods, including views, materialized views, and insert and update statements, are used to store data in the KC Patient Database. Due to the large number of BoCKbase records, especially patient-related data, the enrichment process is very time-consuming.

A second way to enrich patient-related data with BoCKs is directly driven by the users while curating a case in the KC. Curators can create new BoCKs for molecular biomarkers identified in patient samples at the level of integrated biomarkers, genetic biomarkers, or recommendations. These entries are stored in the BoCKbase and labeled as KC-generated entries. After review by an independent expert curation team, the flag is removed, and the new entries become full BoCKs accessible to other KC users. By keeping patient data and BoCKs separate, the KC is also suitable in a multisite setting where a shared BoCKbase is maintained and used (**Extended Data Fig. 5**).

### Generation and storage of therapy recommendations

The result of the KC-based curation process is a set of clinical recommendations presented to the MTB along with the underlying biomarkers and supporting BoCKs. These recommendations are stored in the KC Patient Database, not as individual SQL parameters but as a case-specific JSON file containing all information required to describe a specific recommendation.

### KC backend and frontend

The KC backend and frontend are programmed in Java and Jakarta Server Faces. The PrimeFaces framework is used for the frontend. The ranked mRNA expression is visualized in an R-based Shiny app.

### Public KC instance

The KC and the five use cases presented can be explored in a public instance, which can be accessed by opening an account at registration.public.kc.dkfz.de. The account will be active for four weeks, and the number of simultaneous users can be limited. The public instance has limitations compared to the one in productive use at DKFZ since February 2022. The BoCKbase contains no data from or links to commercial software or databases. All other external knowledge bases are current as of April 1, 2024, and have not been regularly updated. Only selected, representative BoCKs related to the biomarkers described in the use cases are included. The public KC instance provides all functions essential for MTB preparation, e.g., for creating BoCKs or curating and documenting recommendations. To prevent the public instance from deviating from its original state due to external entries, the KC Database is reset to its original state every Sunday at 1 pm Central European Time.

## Data Availability

The Knowledge Connector (KC) is a web application to support MTB workflows, which use multiomics for clinical decision-making. A demo instance is available at https://demo.kc.dkfz.de. In order to comply with requirements for the protection of patient information, this demo instance contains only mock patients. The data of these patients is available at https://demo.kc.dkfz.de. Notably, as this work does not present a clinical trial, aspects of randomization, control group, blinding, or power analysis are not applicable. The code is available at https://git.dkfz.de/cto-sudo-pub/demo.kc.

https://demo.kc.dkfz.de/

---

**Extended Data Fig. 1:**
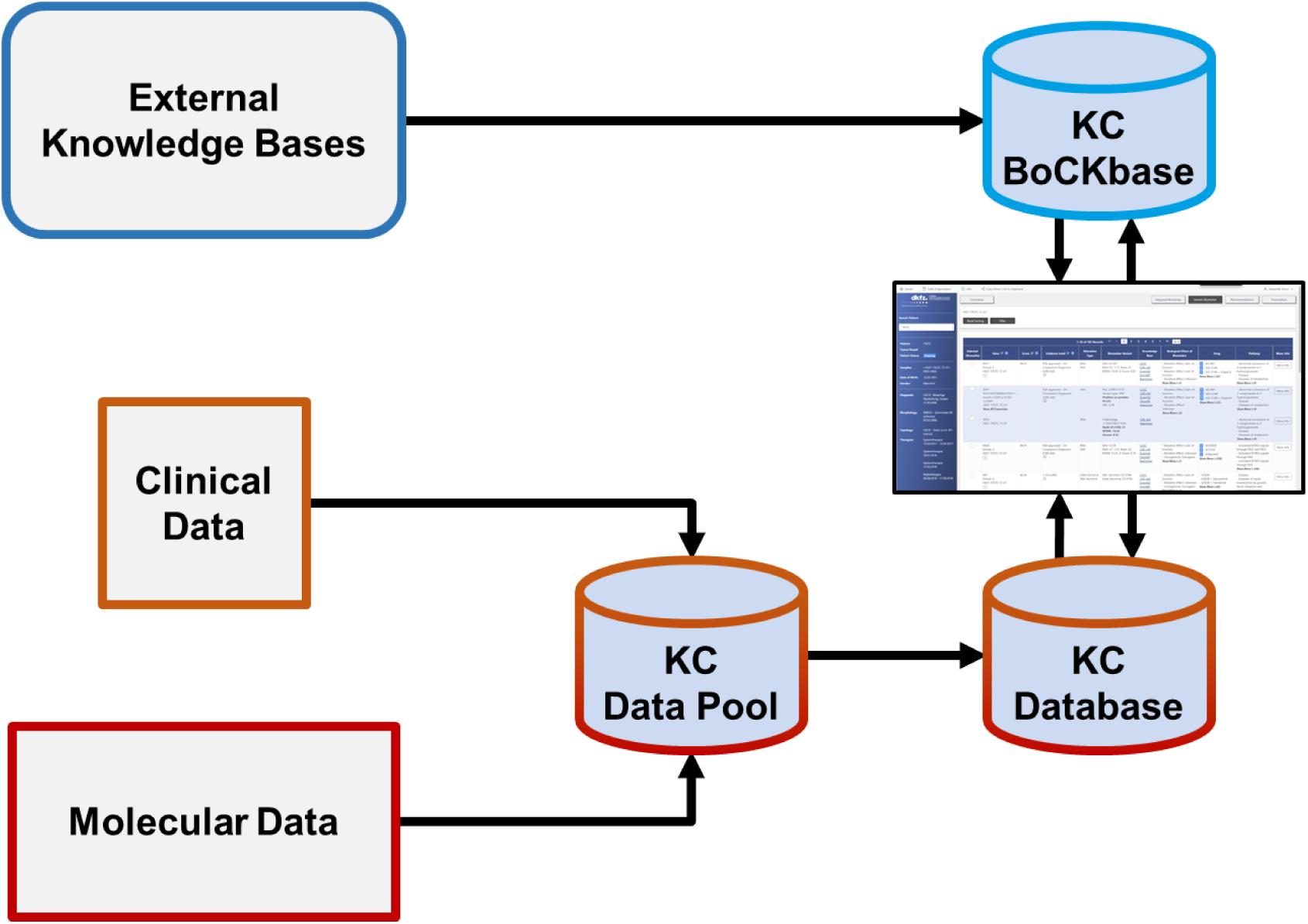
Data flow of the KC. Data from external knowledge bases are extracted and imported into the BoCKbase. Patient-related clinical and molecular data from EHRs and the bioinformatics workflow are stored in the KC Data Pool in a defined data model, prepared for display in the KC, and transferred to the KC Database. In the KC, the BoCKbase is queried for relevant information based on the patient data.

**Extended Data Fig. 2:**
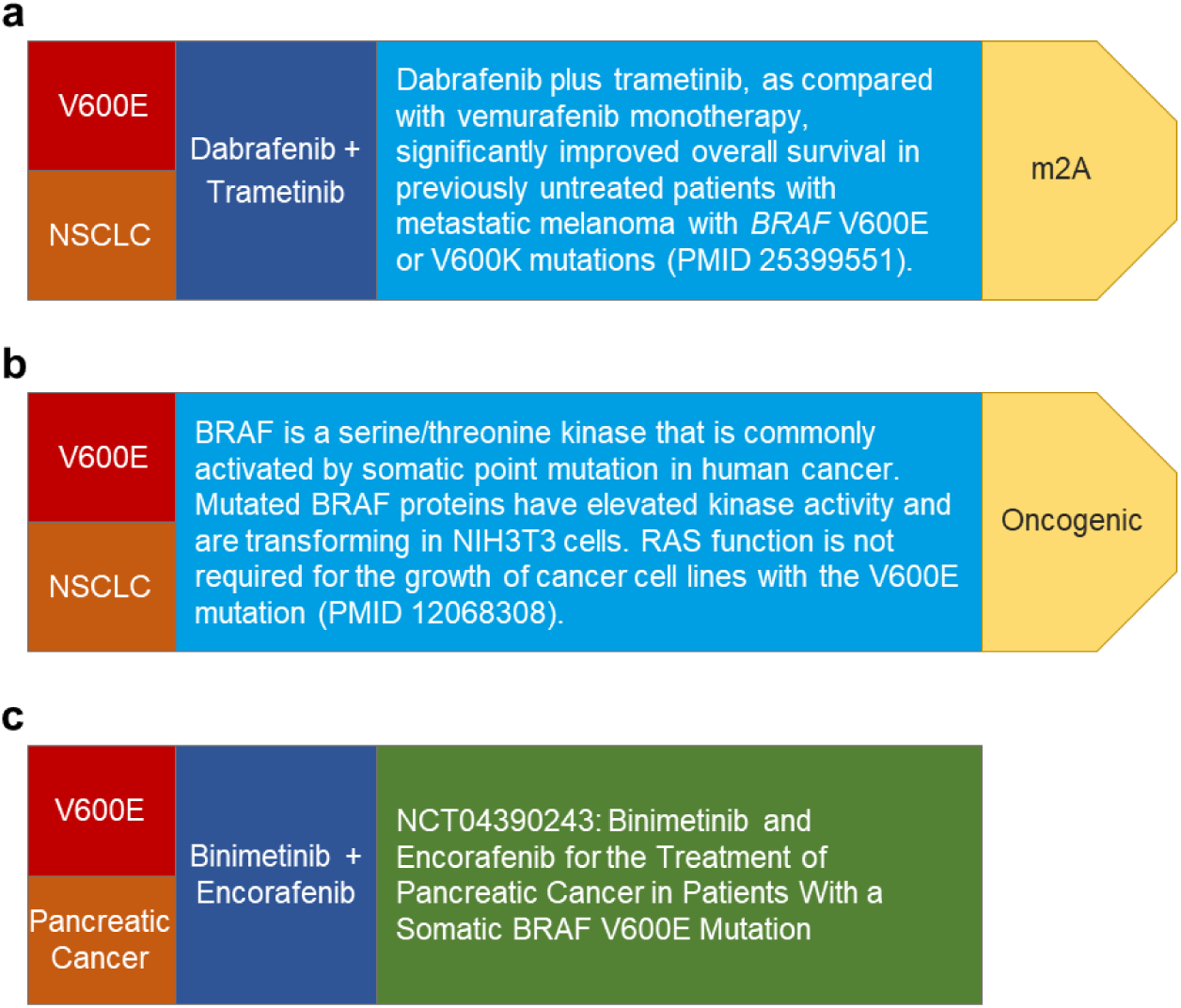
Examples of BoCKs. **a,** BoCK linking a BRAF V600E mutation with dabrafenib and trametinib combination therapy in non-small cell lung cancer (NSCLC) and assigning a predictive statement with a specific mEL. **b,** BoCK linking a BRAF V600E mutation with the functional assessment of its oncogenicity in NSCLC. **c,** BoCK combining a BRAF V600E mutation with eligibility for a clinical trial with encorafenib and binimetinib.

**Extended Data Fig. 3:**
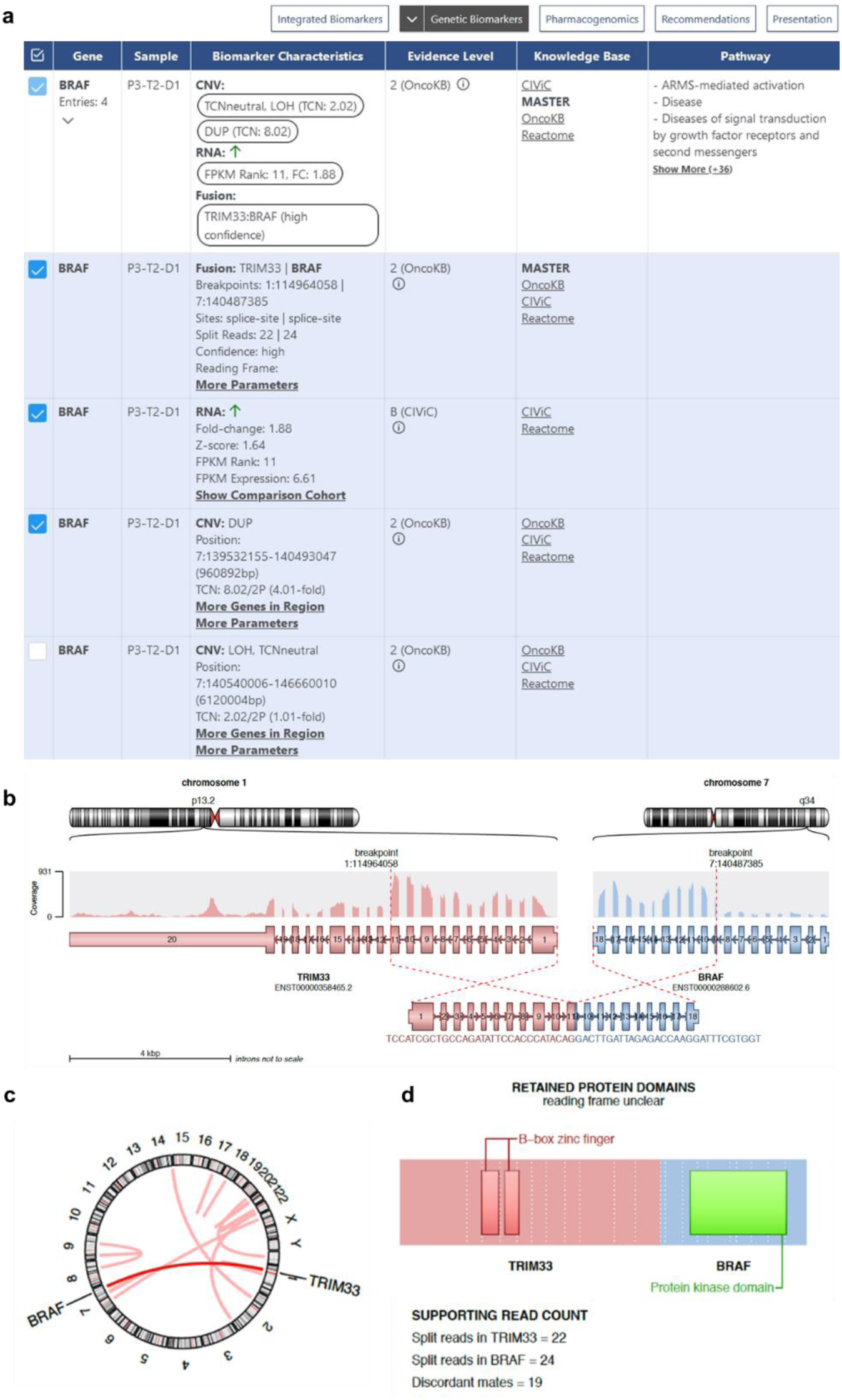
KC display of a TRIM33::BRAF fusion. **a,** Screenshot of the “Genetic Biomarkers” panel illustrating the congruent detection of the fusion gene and an overexpressed fusion transcript. **b,** Visualization of the TRIM33::BRAF fusion event by the fusion detection tool Arriba. **c,** Chromosomal position of the TRIM33::BRAF fusion. **d,** Domain structure of the resulting TRIM33::BRAF fusion protein.

**Extended Data Fig. 4:**
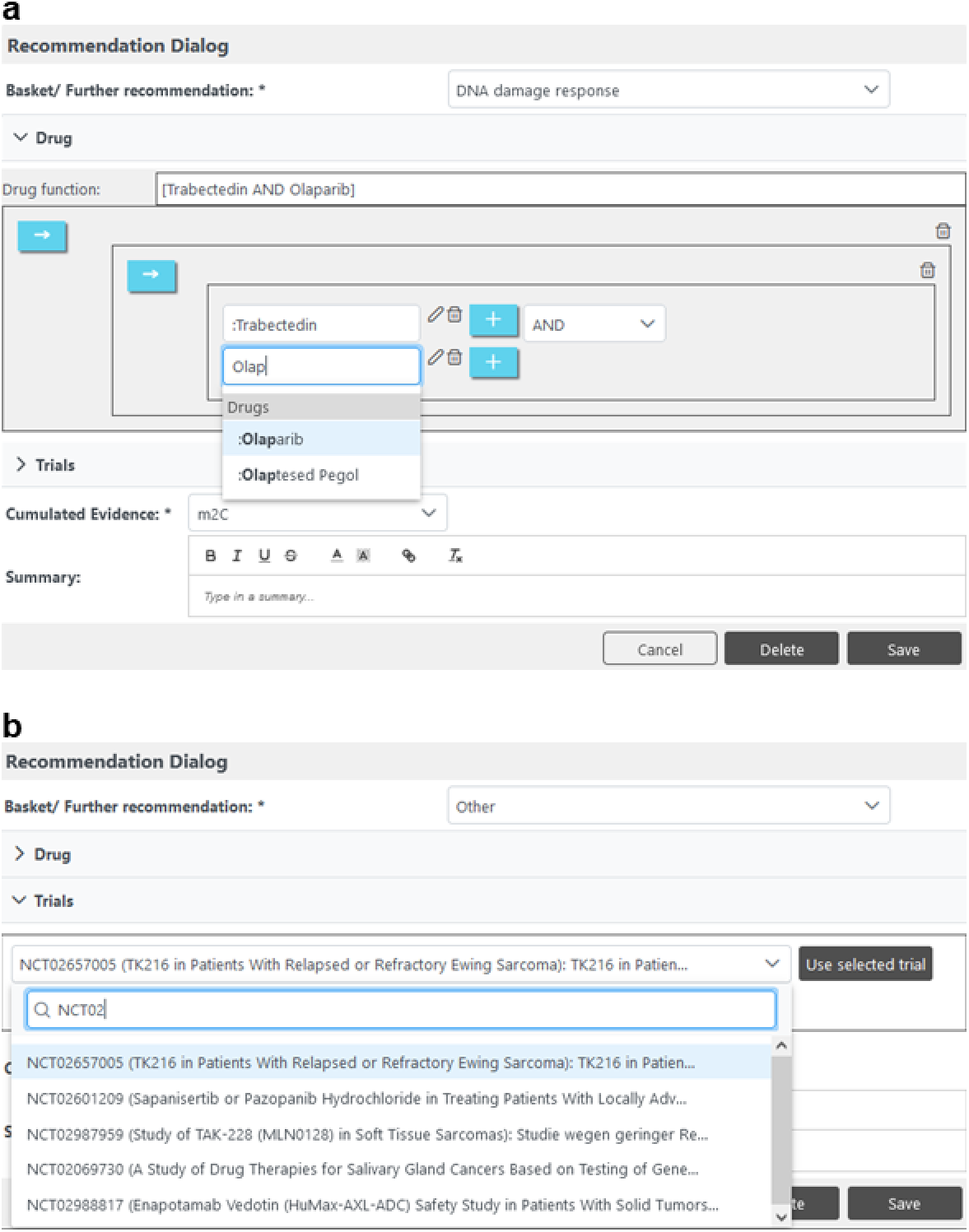
Drug and clinical trial assignment to MTB recommendations. **a,** Recommendation dialog illustrating the assignment of single or multiple drugs to individual treatment recommendations. **b,** Recommendation dialog illustrating the search for and assignment of clinical trial BoCKs to individual treatment recommendations.

**Extended Data Fig. 5:**
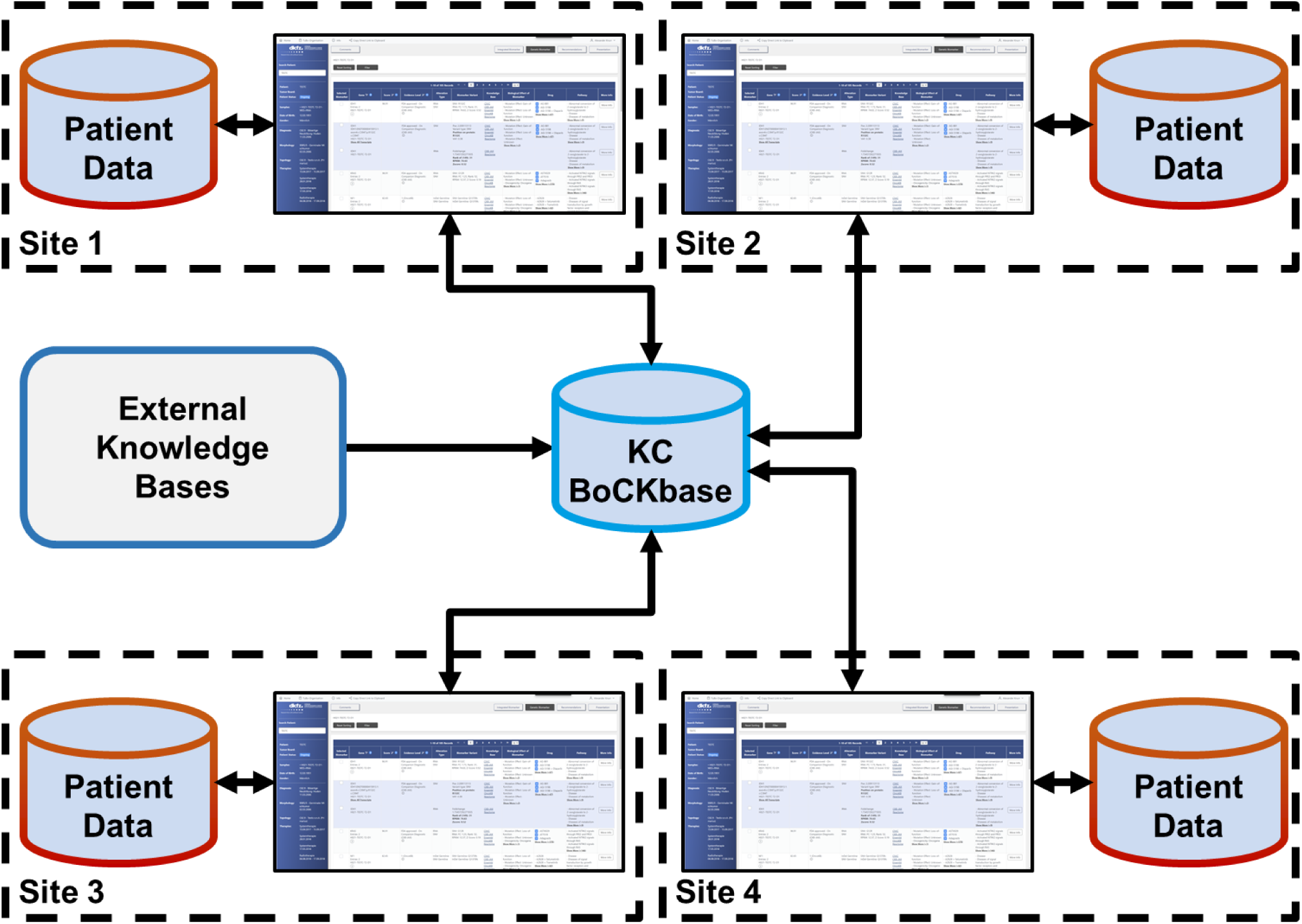
Cross-site data flows of the KC. By separating the KC BoCKbase from the KC Data Pool and the KC Database, a joint BoCKbase can be created and used by collaborating institutions while the patient data remain at the individual sites.

