## Supplemental Data for "Knowledge Connector: Decision support system for multiomics-based precision oncology"

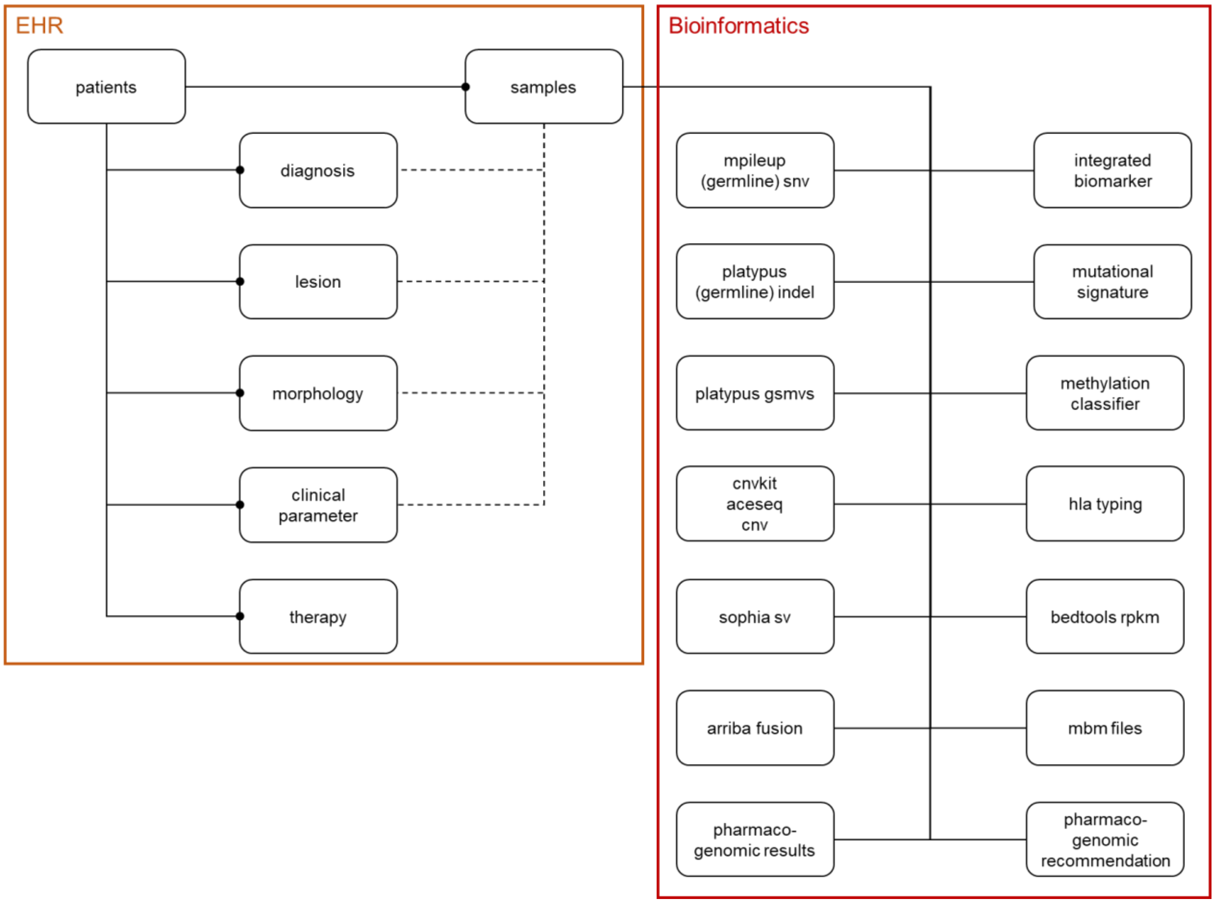


**Supplementary Fig. 1: Patient-related data elements stored in a defined data model in the KC Data Pool.**
Clinical data from EHRs are linked to the patient and can optionally be linked to the corresponding sample. The bioinformatics workflow provides quality-filtered and annotated molecular data linked to the patient via the sample.

**Supplementary Table 1: Patient data from EHRs**

| Column | Type | Description |
|---|---|---|
| **pid** | varchar(32) | Patient identifier |
| **gender** | varchar(16) | Gender |
| **first_name** | varchar(64) | First name |
| **last_name** | varchar(64) | Last name |
| **birthdate** | date | Date of birth |
| **site** | varchar(128) | Enrolling site |
| **applicant** | varchar(128) | Referring physician |

**Supplementary Table 2: Diagnostic data from EHRs**

| Column | Type | Description |
|---|---|---|
| **pid** | varchar(32) | Patient identifier |
| **sample** | varchar(64) | Biosample identifier |
| **diagnosis_type** | varchar(32) | Classification system for diagnosis (e.g., ICD-10) |
| **diagnosis_value** | varchar(256) | Diagnosis (e.g., C50.1) |
| **entity** | varchar(256) | Entity (e.g., non-small cell lung cancer) |
| **documentation_date** | timestamp | Date of documentation |

**Supplementary Table 3: Tumor manifestation data from EHRs**

| Column | Type | Description |
|---|---|---|
| **pid** | varchar(32) | Patient identifier |
| **sample** | varchar(64) | Biosample identifier |
| **lesion** | varchar(256) | ICD-O-3 topology |
| **lesion_type** | varchar(256) | Primary tumor or metastasis |
| **documentation_date** | timestamp | Date of documentation |

**Supplementary Table 4: Tumor morphology data from EHRs**

| Column | Type | Description |
|---|---|---|
| **pid** | varchar(32) | Patient identifier |
| **sample** | varchar(64) | Biosample identifier |
| **morphology_type** | varchar(32) | Classification system for morphology (e.g., ICD-O-3) |
| **morphology_value** | varchar(256) | Morphology (e.g., 8500/3) |
| **tumor_content** | varchar(256) | Tumor cell content |
| **grading** | varchar(256) | Histologic grading |
| **documentation_date** | timestamp | Date of documentation |

**Supplementary Table 5: Clinical parameters from EHRs**

| Column | Type | Description |
|---|---|---|
| **pid** | varchar(32) | Patient identifier |
| **sample** | varchar(64) | Biosample identifier |
| **type** | varchar(32) | Parameter (e.g., ECOG performance status) |
| **value** | varchar(128) | Value (e.g., 1) |
| **documentation_date** | timestamp | Date of documentation |

**Supplementary Table 6: Treatment data from EHRs**

| Column | Type | Description |
|---|---|---|
| **pid** | varchar(32) | Patient identifier |
| **sample** | varchar(64) | Biosample identifier |
| **therapy** | varchar(256) | Free text description of treatment |
| **therapy_start** | timestamp | Start of treatment |
| **therapy_end** | timestamp | End of treatment |
| **cycles** | varchar(256) | Treatment cycles |
| **response** | varchar(256) | Treatment response |
| **best_response** | varchar(256) | Best treatment response |
| **best_response_start** | timestamp | Start of best treatment response |
| **best_response_end** | timestamp | End of best treatment response |

**Supplementary Table 7: Tumor sample data from EHRs**

| Column | Type | Description |
|---|---|---|
| **sample** | varchar(64) | Biosample identifier |
| **pid** | varchar(32) | Patient identifier |
| **sequencing** | varchar(64) | Sequencing method (e.g., WGS) |
| **sequenced** | boolean | Availability of sequencing data available (yes or no) |
| **documentation_date** | timestamp | Date of documentation |

**Supplementary Table 8: Integrated biomarkers data from bioinformatics workflow**

| Column | Type | Description |
|---|---|---|
| **pid** | varchar(32) | Patient identifier |
| **sample** | varchar(64) | Biosample identifier |
| **diagnosis** | varchar(256) | Free text description of diagnosis |
| **hrd** | integer | HRD score |
| **lst** | integer | LST score |
| **tai** | integer | TAI score |
| **snvs** | integer | Number of functional somatic SNVs |
| **msisensor** | numeric | MSIsensor score |
| **ploidy** | integer | Base ploidy where most segments are localized |
| **purity** | integer | Tumor cell content |
| **indels** | integer | Number of functional somatic indels |
| **signatures** | varchar(2048) | Mutational signatures found |
| **somatic** | varchar(2048) | Genes with somatic alterations relevant for inclusion in the NCT PMO-1603/TOP-ART trial (ClinicalTrials.gov: NCT03127215) |
| **germline** | varchar(2048) | Genes with germline alterations relevant for inclusion in the NCT PMO-1603/TOP-ART trial |
| **cellcycle** | varchar(2048) | Biomarkers used for the “Cell Cycle” basket of the DKFZ/NCT/DKTK MASTER program |
| **ras** | varchar(2048) | Biomarkers used for the “RAF-MEK-ERK” basket of the DKFZ/NCT/DKTK MASTER program |
| **rtk** | varchar(2048) | Biomarkers used for the “Tyrosine Kinase” basket of the DKFZ/NCT/DKTK MASTER program |
| **developmental** | varchar(2048) | Biomarkers used for the “Developmental Pathways” basket of the DKFZ/NCT/DKTK MASTER program |
| **mtor** | varchar(2048) | Biomarkers used for the “PI3K-AKT-mTOR” basket of the DKFZ/NCT/DKTK MASTER program |

| | | |
|---|---|---|
| **dnarepair** | varchar(2048) | Biomarkers used for the “DNA Damage Response” basket of the DKFZ/NCT/DKTK MASTER program |
| **immunotherapy** | varchar(2048) | Biomarkers used for the “Immune Evasion” basket of the DKFZ/NCT/DKTK MASTER program |
| **other** | varchar(2048) | Biomarkers used for the “Other” basket of the DKFZ/NCT/DKTK MASTER program |
| **sequencing** | varchar(256) | Sequencing method (e.g., WGS) |
| **tmb** | numeric | Tumor mutational burden |
| **tmbhigh** | numeric | Cut-off for the definition of high tumor mutational burden (unit: non-synonymous mutations per coding megabase) |
| **telomerecontent** | numeric | Telomere content ratio |
| **altscore** | numeric | Alternative lengthening of telomeres score |
| **baskets_geneswithhits** | varchar(256) | List of genes from the DKFZ/NCT/DKTK MASTER baskets with alterations |
| **topart_geneswithhits** | varchar(256) | List of genes with alterations relevant for inclusion in the NCT PMO-1603/TOP-ART trial |
| **quality** | varchar(256) | Quality of RNA-seq data |
| **documentation_date** | timestamp | Date of documentation |

**Supplementary Table 9: Mutational signatures data from bioinformatics workflow**

| Column | Type | Description |
|---|---|---|
| **pid** | varchar(32) | Patient identifier |
| **sample** | varchar(64) | Biosample identifier |
| **mutational_signature _sample** | varchar(64) | Set of mutational signatures (Valid: validated signatures; Artif: including artifact signatures) and algorithm (gen: general; abs: absolute, used for WGS; norm: normalized, used for WES) |
| **mutational_signature** | varchar(16) | Mutational signature |
| **normalised_upper _border** | numeric | Upper border of normalized exposure |
| **normalised_lower _border** | numeric | Lower border of normalized exposure |
| **relative_upper_border** | numeric | Relative upper border of exposure |
| **relative_lower_border** | numeric | Relative lower border of exposure |
| **normalised_exposure** | numeric | Normalized exposure |
| **exposure** | numeric | Exposure |
| **upper_border** | numeric | Upper border of exposure |
| **lower_border** | numeric | Lower border of exposure |
| **documentation_date** | timestamp | Date of documentation |

**Supplementary Table 10: HLA typing data from bioinformatics workflow**

| Column | Type | Description |
|---|---|---|
| **pid** | varchar(32) | Patient identifier |
| **sample** | varchar(64) | Biosample identifier |
| **hla_locus** | varchar(64) | HLA gene |
| **hla_sample_name** | varchar(64) | Sample type the HLA type was called from (control, tumor; DNA, RNA) |
| **hla_sample_value** | varchar(64) | HLA allele |
| **documentation_date** | timestamp | Date of documentation |

**Supplementary Table 11: Methylation classifier data from bioinformatics workflow**

| Column | Type | Description |
|---|---|---|
| **pid** | varchar(32) | Patient identifier |
| **sample** | varchar(64) | Biosample identifier |
| **type** | varchar(64) | Classifier type |
| **version** | varchar(64) | Version of classifier type |
| **prediction** | varchar(256) | Code of predicted diagnosis |
| **probability** | numeric | Probability of matching diagnosis |
| **diagnosis** | varchar(256) | Full text description of predicted diagnosis |
| **description** | varchar(2048) | Description of diagnosis cluster |
| **documentation_date** | timestamp | Date of documentation |

**Supplementary Table 12: Somatic SNV data from the somatic SNV calling workflow**

| Column | Type | Description |
|---|---|---|
| **pid** | varchar(32) | Patient identifier |
| **sample** | varchar(64) | Biosample identifier |
| **chromosome** | varchar(128) | Chromosome |
| **position** | integer | 1-based position of variant |
| **dbsnp** | varchar(4096) | dbSNP rs identifier |
| **base_ref** | varchar(256) | Reference allele |
| **base_alt** | varchar(256) | Alternative allele |
| **confidence** | integer | Quality score associated with alleles inferred |
| **filter** | varchar(256) | Flag indicating which filters variant has failed or PASS if all filters were passed |
| **genes** | varchar(128) | HUGO gene symbols |
| **tumor_dna_af** | numeric | Variant allele frequency (tumor DNA) |
| **tumor_rna_af** | numeric | Variant allele frequency (tumor RNA) |
| **control_dna_af** | numeric | Variant allele frequency (control DNA) |
| **exonic_classification** | varchar(256) | Impact of variant on protein translation |
| **reclassification** | varchar(256) | Classification of variant as somatic or germline |
| **annovar_transcripts** | varchar(4096) | Information on transcripts annotated by ANNOVAR |
| **annovar_function** | varchar(4096) | Location of variant (intron, exon, splice site, untranslated region, upstream, downstream) |
| **ref_genome** | varchar(16) | Reference genome used |
| **documentation_date** | timestamp | Date of documentation |

**Supplementary Table 13: Germline SNV data from the somatic SNV calling workflow**

| Column | Type | Description |
|---|---|---|
| **pid** | varchar(32) | Patient identifier |
| **sample** | varchar(64) | Biosample identifier |
| **chromosome** | varchar(128) | Chromosome |
| **position** | integer | 1-based position of variant |
| **dbsnp** | varchar(4096) | dbSNP rs identifier |
| **base_ref** | varchar(256) | Reference allele |
| **base_alt** | varchar(256) | Alternative allele |
| **confidence** | integer | Quality score associated with alleles inferred |
| **filter** | varchar(256) | Flag indicating which filters variant has failed or PASS if all filters were passed |
| **genes** | varchar(128) | HUGO gene symbols |
| **tumor_dna_af** | numeric | Variant allele frequency (tumor DNA) |
| **tumor_rna_af** | numeric | Variant allele frequency (tumor RNA) |
| **control_dna_af** | numeric | Variant allele frequency (control DNA) |
| **exonic_classification** | varchar(256) | Impact of variant on protein translation |
| **reclassification** | varchar(256) | Classification of variant as somatic or germline |
| **annovar_transcripts** | varchar(4096) | Information on transcripts annotated by ANNOVAR |
| **annovar_function** | varchar(4096) | Location of variant (intron, exon, splice site, untranslated region, upstream, downstream) |
| **ref_genome** | varchar(16) | Reference genome used |
| **documentation_date** | timestamp | Date of documentation |

**Supplementary Table 14: Somatic indel data from the somatic indel calling workflow**

| Column | Type | Description |
|---|---|---|
| **pid** | varchar(32) | Patient identifier |
| **sample** | varchar(64) | Biosample identifier |
| **chromosome** | varchar(128) | Chromosome |
| **position** | integer | 1-based position of variant |
| **dbsnp** | varchar(4096) | dbSNP rs identifier |
| **base_ref** | varchar(256) | Reference allele |
| **base_alt** | varchar(256) | Alternative allele |
| **confidence** | integer | Quality score associated with alleles inferred |
| **filter** | varchar(256) | Flag indicating which filters variant has failed or PASS if all filters were passed |
| **genes** | varchar(128) | HUGO gene symbols |
| **tumor_dna_af** | numeric | Variant allele frequency (tumor DNA) |
| **tumor_rna_af** | numeric | Variant allele frequency (tumor RNA) |
| **control_dna_af** | numeric | Variant allele frequency (control DNA) |
| **exonic_classification** | varchar(256) | Impact of variant on protein translation |
| **reclassification** | varchar(256) | Classification of variant as somatic or germline |
| **annovar_transcripts** | varchar(4096) | Information on transcripts annotated by ANNOVAR |
| **annovar_function** | varchar(4096) | Location of variant (intron, exon, splice site, untranslated region, upstream, downstream) |
| **ref_genome** | varchar(16) | Reference genome used |

| | | |
|---|---|---|
| **documentation_date** | timestamp | Date of documentation |

**Supplementary Table 15: Germline indel data from the somatic indel calling workflow**

| Column | Type | Description |
|---|---|---|
| **pid** | varchar(32) | Patient identifier |
| **sample** | varchar(64) | Biosample identifier |
| **chromosome** | varchar(128) | Chromosome |
| **position** | integer | 1-based position of variant |
| **dbsnp** | varchar(4096) | dbSNP rs identifier |
| **base_ref** | varchar(256) | Reference allele |
| **base_alt** | varchar(256) | Alternative allele |
| **confidence** | integer | Quality score associated with alleles inferred |
| **filter** | varchar(256) | Flag indicating which filters variant has failed or PASS if all filters were passed |
| **genes** | varchar(128) | HUGO gene symbols |
| **tumor_dna_af** | numeric | Variant allele frequency (tumor DNA) |
| **tumor_rna_af** | numeric | Variant allele frequency (tumor RNA) |
| **control_dna_af** | numeric | Variant allele frequency (control DNA) |
| **exonic_classification** | varchar(256) | Impact of variant on protein translation |
| **reclassification** | varchar(256) | Classification of variant as somatic or germline |
| **annovar_transcripts** | varchar(4096) | Information on transcripts annotated by ANNOVAR |
| **annovar_function** | varchar(4096) | Location of variant (intron, exon, splice site, untranslated region, upstream, downstream) |
| **ref_genome** | varchar(16) | Reference genome used |
| **documentation_date** | timestamp | Date of documentation |

**Supplementary Table 16: Germline small variant (i.e., SNV and indel) data from the germline variant calling workflow**

| Column | Type | Description |
|---|---|---|
| **pid** | varchar(32) | Patient identifier |
| **sample** | varchar(64) | Biosample identifier |
| **chromosome** | varchar(128) | Chromosome |
| **position** | integer | 1-based position of variant |
| **base_ref** | varchar(256) | Reference allele |
| **base_alt** | varchar(256) | Alternative allele |
| **filter** | varchar(256) | Flag indicating which filters variant has failed or PASS if all filters were passed |
| **genes** | varchar(128) | HUGO gene symbols |
| **transcript** | varchar(256) | Ensembl transcript ID |
| **variant_on_gene** | varchar(256) | Effect of variant on cDNA according to HGVSc |
| **variant_on_protein** | varchar(256) | Effect of variant on protein according to HGVSp |
| **rna_variant_expression** | varchar(256) | Text string indicating if variant was expressed (tumor RNA) |

| | | |
|---|---|---|
| **tumor_dna_af** | numeric | Variant allele frequency (tumor DNA) |
| **tumor_rna_af** | numeric | Variant allele frequency (tumor RNA) |
| **control_dna_af** | numeric | Variant allele frequency (control DNA) |
| **max_gnom_ad_af** | varchar(256) | Maximum alternative/minor allele frequency in GnomAD |
| **max_gnom_ad_ac** | varchar(256) | Maximum count of individuals carrying the alternative/minor allele in GnomAD |
| **max_gnom_ad_homo** | varchar(256) | Maximum count of individuals carrying the alternative/minor allele homozygously in GnomAD |
| **max_lc_vf** | varchar(256) | Maximum variant frequency in local control WGS and WES datasets |
| **vep_most_ severe_consequence** | varchar(256) | Most severe consequence of variant on coding sequence according to Variant Effect Predictor |
| **all_functional_consensus** | varchar(1028) | Consensus functional consequence on coding sequence |
| **cadd_phred** | varchar(256) | Prediction of variant pathogenicity according to CADD score on PHRED scale |
| **acmg_classification** | varchar(256) | Pathogenicity classification according to ACMG criteria |
| **variant_classification** | varchar(256) | Impact of variant on protein translation |
| **impact** | varchar(256) | Predicted impact of variant |
| **charger_score** | varchar(256) | Pathogenicity score of variant calculated with CharGer |
| **charger_summary** | varchar(4096) | Pathogenicity of variant calculated with CharGer |
| **charger_classification** | varchar(256) | Pathogenicity classification of variant calculated with CharGer |
| **clinvar_pathogenicity** | varchar(256) | Pathogenicity classification of variant according to ClinVar |
| **clinvar_traits** | varchar(4096) | Traits associated with variant in ClinVar |
| **clnid** | varchar(256) | ClinVar identifier |
| **cln_inclusion_status** | varchar(256) | Flag if variant is present in ClinVar inclusion list |
| **clnrevstat** | varchar(256) | Review status of variant in ClinVar |
| **clnsig** | varchar(256) | Pathogenicity of variant in ClinVar |

| | | |
|---|---|---|
| **clnsigconf** | varchar(256) | Confidence of variant pathogenicity assessement in ClinVar |
| **hgnc_url** | varchar(256) | Link to gene entry in HGNC |
| **hgvsg_exon_intron** | varchar(256) | Number of variant-containing exon or intron |
| **splice_ai_ds_gt_04** | varchar(256) | Predicted effect of variant on splicing pattern according to SpliceAI with high recall and low precision (0.4) |
| **splice_ai_ds_gt_09** | varchar(256) | Predicted effect of variant on splicing pattern according to SpliceAI with low recall and high precision (0.9) |
| **tumor_sample** | varchar(256) | Tumor sample used for variant calling |
| **ref_genome** | varchar(16) | Reference genome used |
| **documentation_date** | timestamp | Date of documentation |

#### Supplementary Table 17: CNV data (WES) from bioinformatics workflow

| Column | Type | Description |
|---|---|---|
| **pid** | varchar(32) | Patient identifier |
| **sample** | varchar(64) | Biosample identifier |
| **cnv_type** | varchar(32) | Type of CNV |
| **chromosome** | varchar(128) | Chromosome |
| **start** | integer | Start of CNV |
| **end** | integer | End of CNV |
| **length** | integer | Length of CNV |
| **genes** | varchar(128) | Genes affected by CNV |
| **tcn** | numeric | Total copy number |
| **ploidy** | integer | Base ploidy where most segments are localized |
| **ref_genome** | varchar(16) | Reference genome used |
| **documentation_date** | timestamp | Date of documentation |

#### Supplementary Table 18: CNV data (WGS) from bioinformatics workflow

| Column | Type | Description |
|---|---|---|
| **pid** | varchar(32) | Patient identifier |
| **sample** | varchar(64) | Biosample identifier |
| **cnv_type** | varchar(32) | Type of CNV |
| **chromosome** | varchar(128) | Chromosome |
| **start** | integer | Start of CNV |
| **end** | integer | End of CNV |
| **length** | integer | Length of CNV |
| **genes** | varchar(128) | Genes affected by CNV |
| **bp_genes** | varchar(4096) | Indication if gene located at breakpoint |

| | | |
|---|---|---|
| **tcn** | numeric | Total copy number |
| **ploidy** | integer | Base ploidy where most segments are localized |
| **ref_genome** | varchar(16) | Reference genome used |
| **documentation_date** | timestamp | Date of documentation |

**Supplementary Table 19: SV data from bioinformatics workflow**

| **Column** | **Type** | **Description** |
|---|---|---|
| **pid** | varchar(32) | Patient identifier |
| **sample** | varchar(64) | Biosample identifier |
| **sv_type** | varchar(32) | Type of SV |
| **chromosome_1** | varchar(128) | Information on first chromosome |
| **chromosome_2** | varchar(128) | Information on second chromosome |
| **position_1** | integer | Position of breakpoint on first chromosome |
| **position_2** | integer | Position of breakpoint on second chromosome |
| **event_size** | integer | Distance between breakpoints |
| **event_score** | integer | Score from 1 to 5 indicating reliability of SV call |
| **genes_1** | varchar(128) | HUGO gene symbols |
| **genes_2** | varchar(128) | HUGO gene symbols |
| **direct_fusion _candidates** | varchar(4096) | Potential fusions between genes affected |
| **ref_genome** | varchar(16) | Reference genome used |
| **documentation_date** | timestamp | Date of documentation |

**Supplementary Table 20: Fusion data from bioinformatics workflow**

| Column | Type | Description |
|---|---|---|
| **pid** | varchar(32) | Patient identifier |
| **sample** | varchar(64) | Biosample identifier |
| **pipeline** | varchar(64) | Fusion calling pipeline used |
| **type** | varchar(128) | Type of fusion |
| **confidence** | varchar(64) | Confidence of fusion call |
| **filters** | varchar(256) | Filters used to remove supporting reads |
| **chromosome_1** | varchar(128) | Information on first chromosome |
| **chromosome_2** | varchar(128) | Information on second chromosome |
| **breakpoint_1** | integer | Position of breakpoint on first chromosome |
| **breakpoint_2** | integer | Position of breakpoint on second chromosome |
| **closest_genomic _breakpoint_1** | varchar | DNA breakpoint closest to first RNA breakpoint |
| **closest_genomic _breakpoint_2** | varchar | DNA breakpoint closest to second RNA breakpoint |
| **coverage_1** | integer | Number of reads at first breakpoint |
| **coverage_2** | integer | Number of reads at second breakpoint |
| **direction_1** | varchar(64) | Direction of reads from first fusion partner |
| **direction_2** | varchar(64) | Direction of reads from second fusion partner |
| **site_1** | varchar(128) | Position of functional sites at first breakpoint |
| **site_2** | varchar(128) | Position of functional sites at second breakpoint |
| **split_reads_1** | integer | Number of split reads supporting gene 1 |
| **split_reads_2** | integer | Number of split reads supporting gene 2 |
| **strand_1** | varchar(8) | Transcribed strand for gene 1 |
| **strand_2** | varchar(8) | Transcribed strand for gene 2 |
| **genes_1** | varchar(128) | Gene at fusion N-terminus |

| | | |
|---|---|---|
| **genes_2** | varchar(128) | Gene at fusion C-terminus |
| **transcript_id_1** | varchar(128) | Ensembl transcript identifier for gene 1 |
| **transcript_id_2** | varchar(128) | Ensembl transcript identifier for gene 2 |
| **fusion_transcript** | varchar(2058) | Fusion transcript sequence |
| **discordant_mates** | integer | Number of read pairs of discordant mates supporting fusion |
| **peptide_sequence** | varchar(2058) | Fusion peptide sequence |
| **read_identifiers** | varchar(2058) | Names of supporting reads |
| **reading_frame** | varchar(128) | Reading frame of fusion product |
| **retained_proteine _domains** | varchar(2058) | Protein domains retained by fusion product |
| **tags** | varchar(128) | User-defined tags for positions of interest |
| **ref_genome** | varchar(16) | Reference genome used |
| **documentation_date** | timestamp | Date of documentation |

**Supplementary Table 21: Gene expression data from bioinformatics workflow**

| **Column** | **Type** | **Description** |
|---|---|---|
| **pid** | varchar(32) | Patient identifier |
| **sample** | varchar(64) | Biosample identifier |
| **gene** | varchar(32) | Gene name |
| **expression** | numeric | Expression value (e.g., RPKM, FPKM, TPM) |
| **fold_change** | numeric | Fold-change expression compared to median of reference cohort |
| **z_score** | numeric | z-score of expression value compared to reference cohort |
| **rpkm_rank** | integer | Rank of expression compared to reference cohort |
| **reference_cohort** | varchar(16) | Name of reference cohort |
| **ref_genome** | varchar(16) | Reference genome used |
| **documentation_date** | timestamp | Date of documentation |

**Supplementary Table 22: Multibitmap files from bioinformatics workflow**

| **Column** | **Type** | **Description** |
|---|---|---|
| **pid** | varchar(32) | Patient identifier |
| **sample** | varchar(64) | Biosample identifier |
| **file_data** | bytea | Binary data of any kind (e.g., images, documents) |
| **file_name** | varchar(1024) | File name |
| **file_type** | varchar(64) | File data type |
| **file_date** | timestamp | File date |
| **file_size** | numeric | File size |

| | | |
|---|---|---|
| **documentation_date** | timestamp | Date of file upload |

**Supplementary Table 23: Pharmacogenomics data from bioinformatics workflow**

| Column | Type | Description |
|---|---|---|
| **pid** | varchar(32) | Patient identifier |
| **sample** | varchar(64) | Biosample identifier |
| **gene** | varchar(256) | HUGO gene symbols |
| **harmonized_genotype** | varchar(256) | Consensus genotype from harmonization of genotypes from all tools |
| **harmonized_phenotype** | varchar(256) | Translated phenotype based on harmonized genotype |
| **pharmacogenomic_sample** | varchar(256) | Biosample identifier used by pharmacogenomics pipeline |
| **stargazer_genotype** | varchar(256) | Raw genotype output from Stargazer |
| **stargazer_curated_genotype** | varchar(256) | Curated and formatted genotype from Stargazer |
| **stargazer_genotype_tags** | varchar(256) | Tags with additional information on alleles in Stargazer genotype |
| **aldy_genotype** | varchar(256) | Raw genotype output from aldy |
| **aldy_curated_genotype** | varchar(256) | Curated and formatted genotype from Aldy |
| **aldy_genotype_tags** | varchar(256) | Tags with additional information on alleles in Aldy genotype |
| **pypgx_genotype** | varchar(256) | Raw genotype output from PyPGx |
| **pypgx_curated_genotype** | varchar(256) | Curated and formatted genotype from PyPGx |
| **pypgx_genotype_tags** | varchar(256) | Tags with additional information on alleles in PyPGx genotype |
| **cyrius_genotype** | varchar(256) | Raw genotype output from Cyrius |
| **cyrius_curated_genotype** | varchar(256) | Curated and formatted genotype from Cyrius |
| **cyrius_genotype_tags** | varchar(256) | Tags with additional information on alleles in Cyrius genotype |
| **stargazer_phenotype** | varchar(256) | Translated phenotype based on curated Stargazer genotype |

| | | |
|---|---|---|
| **pypgx_phenotype** | varchar(256) | Translated phenotype based on curated PyPGx genotype |
| **stargazer_dip_sv** | varchar(256) | Additional Stargazer CNV information |
| **pypgx_cnv** | varchar(256) | Additional PyPGx CNV information |
| **harmonization_comment** | varchar(256) | Comments from harmonization process regarding special cases |
| **harm_allele1** | varchar(256) | Allele 1 of harmonized genotype |
| **harm_allele2** | varchar(256) | Allele 2 of harmonized genotype |
| **documentation_date** | timestamp | Date of documentation |

### Supplementary Table 24: Pharmacogenomics recommendations from bioinformatics workflow

| Column | Type | Description |
|---|---|---|
| **pid** | varchar(32) | Patient identifier |
| **sample** | varchar(64) | Biosample identifier |
| **gene** | varchar(256) | HUGO gene symbols |
| **harmonized_genotype** | varchar(256) | Consensus genotype from harmonization of genotypes from all tools |
| **harmonized_phenotype** | varchar(256) | Translated phenotype based on harmonized genotype |
| **pharmacogenomic _sample** | varchar(256) | Biosample identifier used by the pharmacogenomics pipeline |
| **drug** | varchar(1028) | Drug |
| **recommendation** | varchar(4096) | Free text of recommendation provided by knowledge base |
| **knowledgebase** | varchar(256) | Underlying knowledge base (e.g., CPIC) |
| **documentation_date** | timestamp | Date of documentation |

### Supplementary Table 25: Knowledge bases selected for content relevant to clinical decision-making

| Knowledge base | Queries | Transcripts | Oncogenicity | Biological effect | Drugs and therapies | Pathways | Gene description |
|---|---|---|---|---|---|---|---|
| CIViC[A] | Gene, variant | | | | X | | X |
| JAX-CKB[B,C] | Gene, variant | | X | X | X | | X |
| Ensembl[A] | Gene | X | | | | | |
| OncoKB[A] | Gene, variant | | X | X | X | | |
| Reactome[A] | Gene | | | | | X | |

[A] Publicly available knowledge base
[B] Commercial knowledge base
[C] BoCKs from JAX-CKB are not part of the public KC instance.
